# Current evidence for COVID-19 therapies: a systematic literature review

**DOI:** 10.1101/2020.12.18.20248452

**Authors:** Tobias Welte, Lucy J. Ambrose, Gillian C. Sibbring, Shehla Sheikh, Hana Müllerová, Ian Sabir

## Abstract

Effective therapeutic interventions for the treatment and prevention of COVID-19 are urgently needed. A systematic review was conducted to identify clinical trials of pharmacological interventions for COVID-19 published between 1 December 2019 and 14 October 2020. Data regarding efficacy of interventions, in terms of mortality, hospitalisation and need for ventilation, were extracted from identified studies and synthesised qualitatively.

In total, 42 clinical trials were included. Interventions assessed included antiviral, mucolytic, anti-malarial, anti-inflammatory and immunomodulatory therapies. Some reductions in mortality, hospitalisation and need for ventilation were seen with interferons and remdesivir, particularly when administered early, and with the mucolytic drug, bromhexine. Most studies of lopinavir/ritonavir and hydroxychloroquine did not show significant efficacy over standard care/placebo. Dexamethasone significantly reduced mortality, hospitalisation and need for ventilation *versus* standard care, particularly in patients with severe disease. Evidence for other classes of interventions was limited. Many trials had a moderate-to-high risk of bias, particularly in terms of blinding; most were short-term; and some included low patient numbers.

This review highlights the need for well-designed clinical trials of therapeutic interventions for COVID-19 to increase the quality of available evidence. It also emphasises the importance of tailoring interventions to disease stage and severity for maximum efficacy.

## Introduction

COVID-19 is an infectious disease caused by a newly discovered coronavirus, severe acute respiratory syndrome coronavirus 2 (SARS-CoV-2) [1]. The global pandemic caused by COVID-19 is ongoing, with a projected death toll of almost 2.5 million by 1 February 2021 [2].

Although COVID-19 presents primarily as a respiratory tract infection, increasing data have shown the potential for systemic involvement, including cardiovascular, neurological and dermatological manifestations, in patients who present with the disease [3]. The pathophysiological course of COVID-19 has been proposed to comprise three distinct phases [4]. In the early infection phase, the SARS-CoV-2 virus enters epithelial cells in the nasal cavity and multiplies in the upper respiratory tract, with or without pulmonary involvement [1, 4, 5]. The second phase is characterised by localised pulmonary inflammation and the development of viral pneumonia, with or without hypoxia. In a minority of patients, the disease enters a third ‘host response’ phase, which manifests as an extrapulmonary systemic hyperinflammation syndrome, characterised by high levels of pro-inflammatory cytokines and potentially leading to thrombotic complications, viral sepsis and multi-organ failure [1, 4, 6].

Increasing understanding of the disease pathways involved in COVID-19 has highlighted the importance of selecting and implementing treatments appropriate for the disease stage that patients present with [4, 6]. Since the start of the outbreak, global efforts to validate effective therapeutic interventions for COVID-19 have resulted in the identification of many potential candidates and the initiation of thousands of clinical trials of therapies with diverse mechanisms of action [7-9].

As of November 2020, a small number of treatments have received regulatory approval on the basis of promising results (table 1). These include the antiviral, remdesivir; remdesivir in combination with baricitinib, a Janus kinase inhibitor; dexamethasone, a corticosteroid; convalescent plasma; bamlanivimab, a monoclonal antibody therapy; and casirivimab and imdevimab, a cocktail of two monoclonal antibodies [10-13]. Additionally, one vaccine against SARS-CoV-2 has been approved in Russia following results from two non-randomised phase 1/2 trials [14, 15] and several other vaccines are in late-phase clinical trials. Despite these developments, treatment options for COVID-19 remain limited. The precise proportion of patients requiring hospitalisation is challenging to determine, given the uncertain prevalence of infection [16]. However, it is estimated that up to 20% of patients with COVID-19 have an illness severe enough to warrant hospitalisation, which may require intensive care admission and need for respiratory support [17-20]. Consequently, the COVID-19 pandemic has put substantial pressure on healthcare systems worldwide [21-25], and there remains an urgent need for effective therapies for the prevention and treatment of COVID-19.

**TABLE 1.** Currently approved treatments for COVID-19

| Regulatory body | Treatment | Details of approval |
| --- | --- | --- |
| EMA | Remdesivir | Conditional marketing authorisation for the treatment of COVID-19 in adults and adolescents $\geq 12$ years of age with pneumonia who require supplemental oxygen [13] |
| | Dexamethasone | Endorsed by the EMA's human medicine's committee to treat hospitalised adults and adolescents $\geq 12$ years of age (weighing $\geq 40$ kg) with COVID-19 who are receiving respiratory support [13] |
| US FDA | Remdesivir | Approval for the treatment of adult and paediatric patients $\geq 12$ years of age (weighing $\geq 40$ kg) with COVID-19 requiring hospitalisation [11] |
|  | Convalescent plasma | Emergency use authorisation for the treatment of hospitalised patients with COVID-19 [11] |
| | Bamlanivimab | Emergency use authorisation for the treatment of mild-to-moderate COVID-19 in adult and paediatric patients $\geq 12$ years of age (weighing $\geq 40$ kg) with a positive test for SARS-CoV-2 and who are at high risk for progressing to severe COVID-19 and/or hospitalisation [11] |
| | Baricitinib in combination with remdesivir | Emergency use authorisation for the treatment of suspected or laboratory-confirmed COVID-19 in hospitalised adults and paediatric patients $\geq 2$ years of age requiring supplemental oxygen, invasive mechanical ventilation or ECMO [11] |
| | Casirivimab and imdevimab | Emergency use authorisation for the treatment of mild to moderate COVID-19 in adults and paediatric patients ($\geq 12$ years weighing at least 40 kg) with positive results of direct SARS-CoV-2 viral testing, and who are at high risk for progressing to severe COVID-19 and/or hospitalisation [11] |
| Russian Ministry of Health | Adenovirus-based anti-SARS-CoV-2 vaccine (Sputnik V) | Approved by the Russian Ministry of Health, based on data from non-randomised phase I/II clinical trials [14, 15] |

COVID-19: Coronavirus disease 2019; ECMO, extracorporeal membrane oxygenation; EMA: European Medicines Agency; US FDA: United States Food and Drug Administration; SARS-CoV-2: severe acute respiratory syndrome coronavirus 2.

The large quantity of clinical data being generated, wide spectrum of disease presentations and rapidly changing clinical landscape present a critical need for comprehensive evidence summaries and treatment comparisons. The overall aim of this systematic literature review (SLR) was to assess available evidence regarding efficacy and safety of potential pharmacological interventions for COVID-19.

Evidence retrieved was considered in the context of the evolving understanding of the pathophysiology of COVID-19, the burden of COVID-19 in terms of mortality and healthcare resource utilisation, and current knowledge gaps. Thus, the aim of the analysis presented here was to synthesise evidence for mortality, hospitalisation and need for ventilation with current therapies.

## Methods

### Overview

This SLR (registered with the Research Registry, unique identifying number: reviewregistry1019) evaluated studies of pharmacological options for the treatment and prevention of COVID-19. The review was conducted according to the principles embodied in the Cochrane Handbook for Systematic Reviews of Interventions [26] and guidance published by the Centre for Reviews and Dissemination [27].

### Search strategy

The systematic literature search was conducted from 1 December 2019 to 14 July 2020 and updated on 14 October 2020, using the electronic databases Embase, MEDLINE^®^ via the PubMed platform and The Cochrane Library. Details of the search strings used for each database are presented in supplementary file 1. Searches were supplemented by review of reports of pharmacological interventions for COVID-19 included in a systematic and living map of COVID-19 evidence [28]. These articles were assessed according to the same eligibility criteria as for the systematic searches. As part of the 14 October update, an additional search of PubMed Central was conducted following retrieval of one article from the systematic and living map that was available in PubMed Central but not in the other databases searched.

### Eligibility criteria

Key inclusion criteria were clinical trials of any pharmacological preventive or treatment approach for COVID-19 of any stage, conducted in human subjects of any age.

Exclusion criteria included studies of non-pharmacological interventions; traditional or herbal medicines; studies that reported on *in vitro* or *in silico* investigations; guidelines; clinical trial protocols or projection studies; or observational studies, such as prospective and retrospective cohort studies, case-control studies, cross-sectional studies, case reports and case series. Articles that were not written in English, reviews, comments, editorials, congress abstracts, and articles that had not undergone peer review were also excluded.

### Study selection

After removal of duplicates, records identified in the electronic database searches were manually screened for eligibility on the basis of titles and abstracts by a single reviewer, with a second reviewer screening ≥10% of records selected at random. Any disagreements or uncertainties were discussed with a third reviewer to achieve a consensus. Subsequently, full texts of potentially relevant studies were obtained and reviewed for eligibility as for the first-pass screening. Studies containing duplicate information or not meeting the inclusion criteria upon further review were excluded.

### Data extraction

Details of study design and population, interventions, comparators, follow-up duration and safety and efficacy outcomes were extracted from identified articles using a pilot-tested data extraction spreadsheet constructed in Microsoft Excel^®^. If available, information on statistical comparisons between interventions was recorded; any studies reporting insufficient data meeting the inclusion criteria were excluded at the data extraction stage. All extracted data were cross-checked by an independent reviewer.

### Data synthesis and quality assessment

Included studies were reviewed and assessed for comparability in terms of study design and outcomes reported. Randomised studies were included in the qualitative synthesis if they reported ≥1 of the selected outcomes: mortality, hospitalisation (any reported outcome, including duration, proportion of patients discharged and incidence) and need for ventilation (use of oxygen, non-invasive ventilation and intensive mechanical ventilation). Non-randomised trials reporting these outcomes were only included if no randomised trial evidence was available for a particular intervention.

Studies were grouped according to intervention and the disease phase targeted by the intervention; details of the categorisation used are presented in supplementary table 1. Studies reporting similar outcome measures were summarised descriptively according to the type of intervention. Studies with outcomes that were defined differently to other studies (*e*.*g*. event-free survival rather than percentage mortality, or outcomes reported for the overall population rather than according to treatment with [or allocation to] specific interventions) could not be grouped, and these were assessed separately.

Each study was assessed in terms of methodological quality based on criteria consistent with those in the Cochrane Handbook for Systematic Reviews of Interventions and the Cochrane risk-of-bias tool [26, 29].

## Results

### Summary of studies included

The literature searches identified 33,674 unique citations, and from these, 436 full-text articles were assessed for eligibility (supplementary figure 1). A total of 375 articles were excluded as they did not meet eligibility criteria for reasons such as reporting no outcomes of interest, presenting duplicate data from other studies or having a non-interventional study design. A total of 61 articles were retained for inclusion in the SLR and, of these, 43 articles, reporting on 42 trials [30-72], were selected for qualitative synthesis, based on whether they reported ≥1 of the outcomes of interest.

**FIGURE 1.**
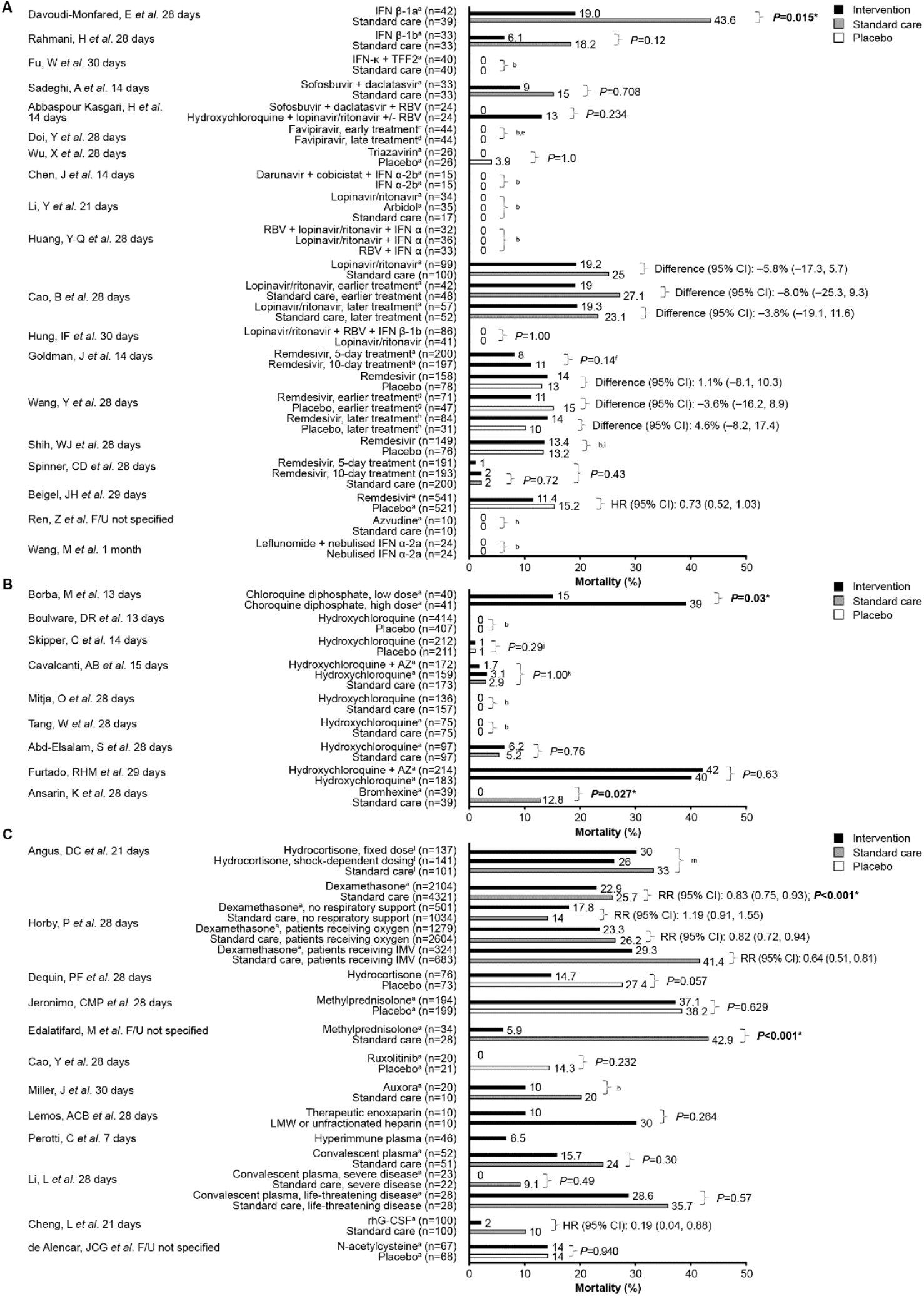
Summary of mortality outcomes in trials of antivirals (A), anti-malarial and mucolytic drugs (B) and other therapies (C) included in the qualitative synthesis. Other therapies include anti-inflammatory drugs, anticoagulants, kinase inhibitors, CRAC channel inhibitors, anticoagulants, immunomodulatory therapies and repair therapies. *: Indicates a statistically significant *P*-value. Results from one study are not presented graphically. Deftereos *et al*. [38] reported event-free survival as a primary outcome, which was defined as survival without meeting the primary clinical endpoint (deterioration by 2 points on a 7-grade clinical status scale, ranging from able to resume normal activities to death). ^a^: Treatment administered in addition to standard care, as defined by the investigators in each trial; ^b^: No between-group comparison; ^c^: Treatment on day 1 of study participation; ^d^: Treatment on day 6 of study participation; ^e^: Outcome was disease progression or death; ^f^: *P*-value for comparison of 7-level ordinal outcome (including death) at 14 days; ^g^: Treatment ≤10 days of symptom onset; ^h^: Treatment >10 days of symptom onset; ^i^: Re-analysis of data from Wang Y, *et al*. using different criteria; ^j^: Outcome was incidence of hospitalisation or death; ^k^: *P*-value for comparisons of the 7-level ordinal outcome (including death) at 15 days; ^l^: Participants could be randomly assigned to other interventions within other therapeutic domains; ^m^: Median (95% CI) adjusted odds ratios *versus* the no-hydrocortisone group were 1.03 (0.53, 1.95) and 1.10 (0.58, 2.11) for the fixed-dose and shock-dependent dosing hydrocortisone groups, respectively. These yielded 54% and 62% Bayesian posterior probabilities of superiority. AZ: azithromycin; CI: confidence interval; CRAC: calcium release-activated calcium; F/U: follow-up; HR: hazard ratio; IFN: interferon; rhG-CSF: recombinant human granulocyte colony-stimulating factor; RR: rate ratio; RBV: ribavirin; TFF2: trefoil factor 2.

Details of the 18 articles that were not retained for qualitative synthesis are summarised in supplementary table 2. An overview of the 43 articles included in the qualitative synthesis is presented in supplementary figure 2, and detailed characteristics of the individual trials are presented in supplementary table 3. The majority of trials were conducted in patients hospitalised with COVID-19, but three trials included non-hospitalised patients [37, 42, 44] and one was conducted in asymptomatic adults with occupational or household exposure to COVID-19 [32] (supplementary figure 2). Most trials used either standard care, which differed between trials, or placebo as a comparator.

**TABLE 2.**
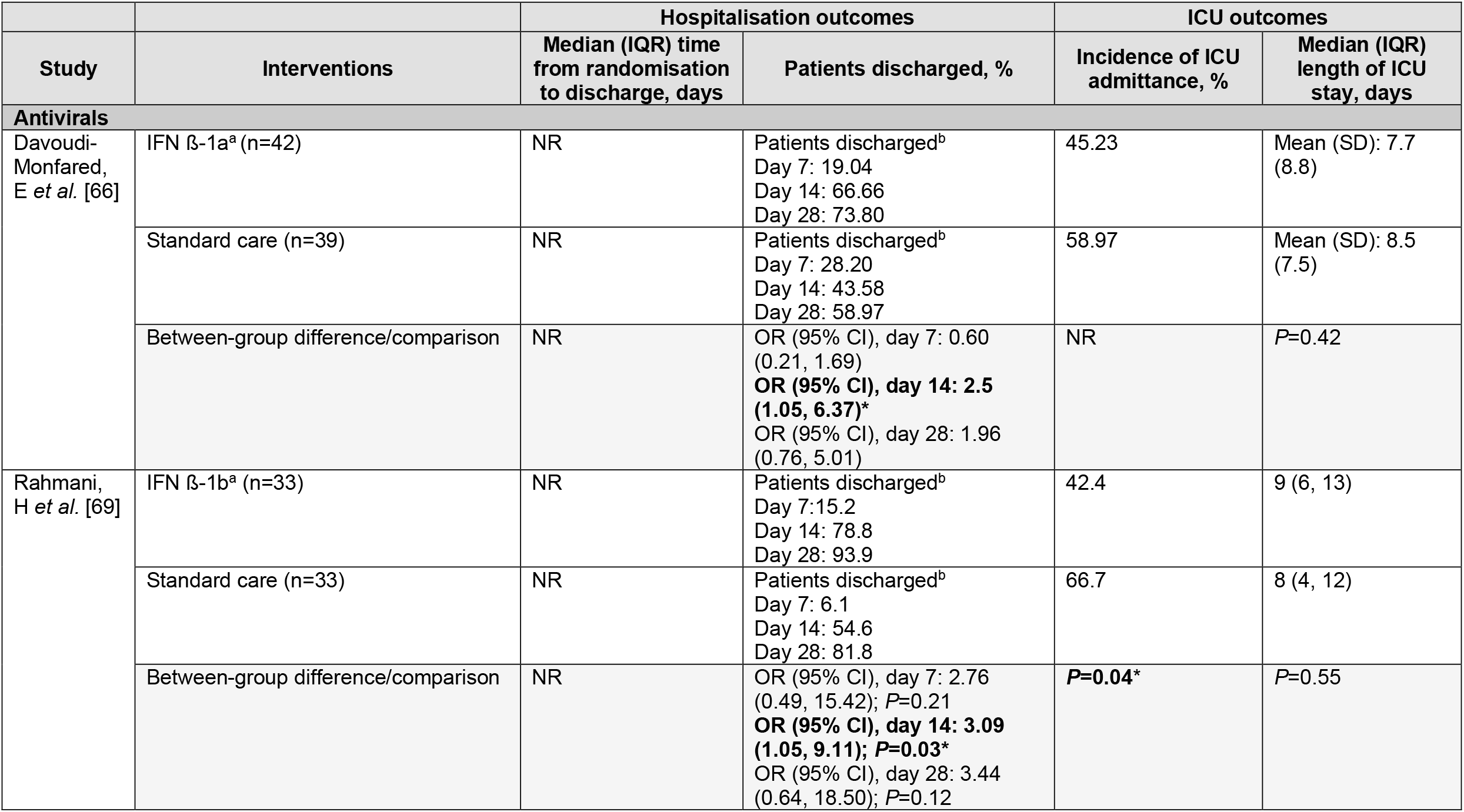
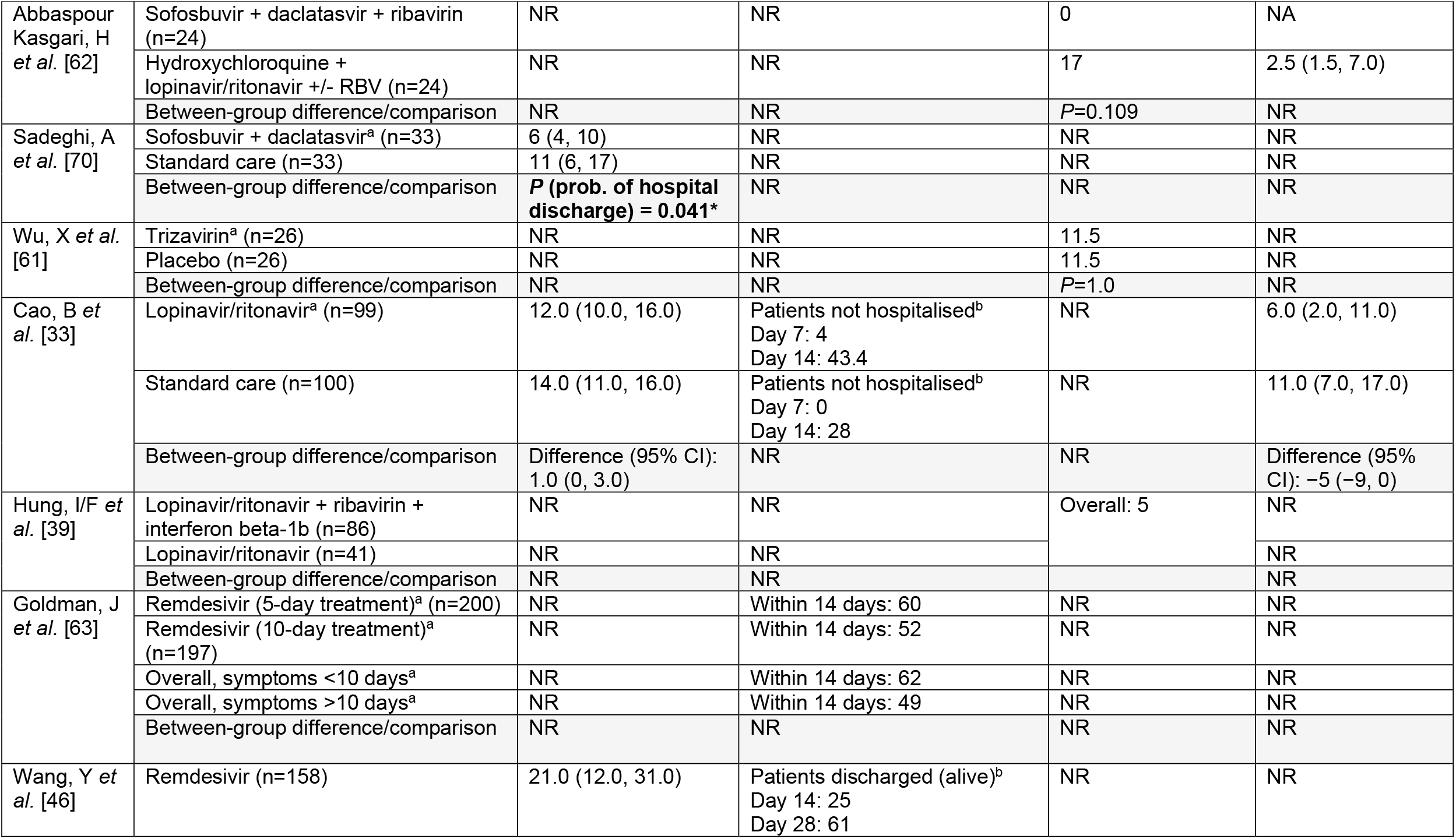
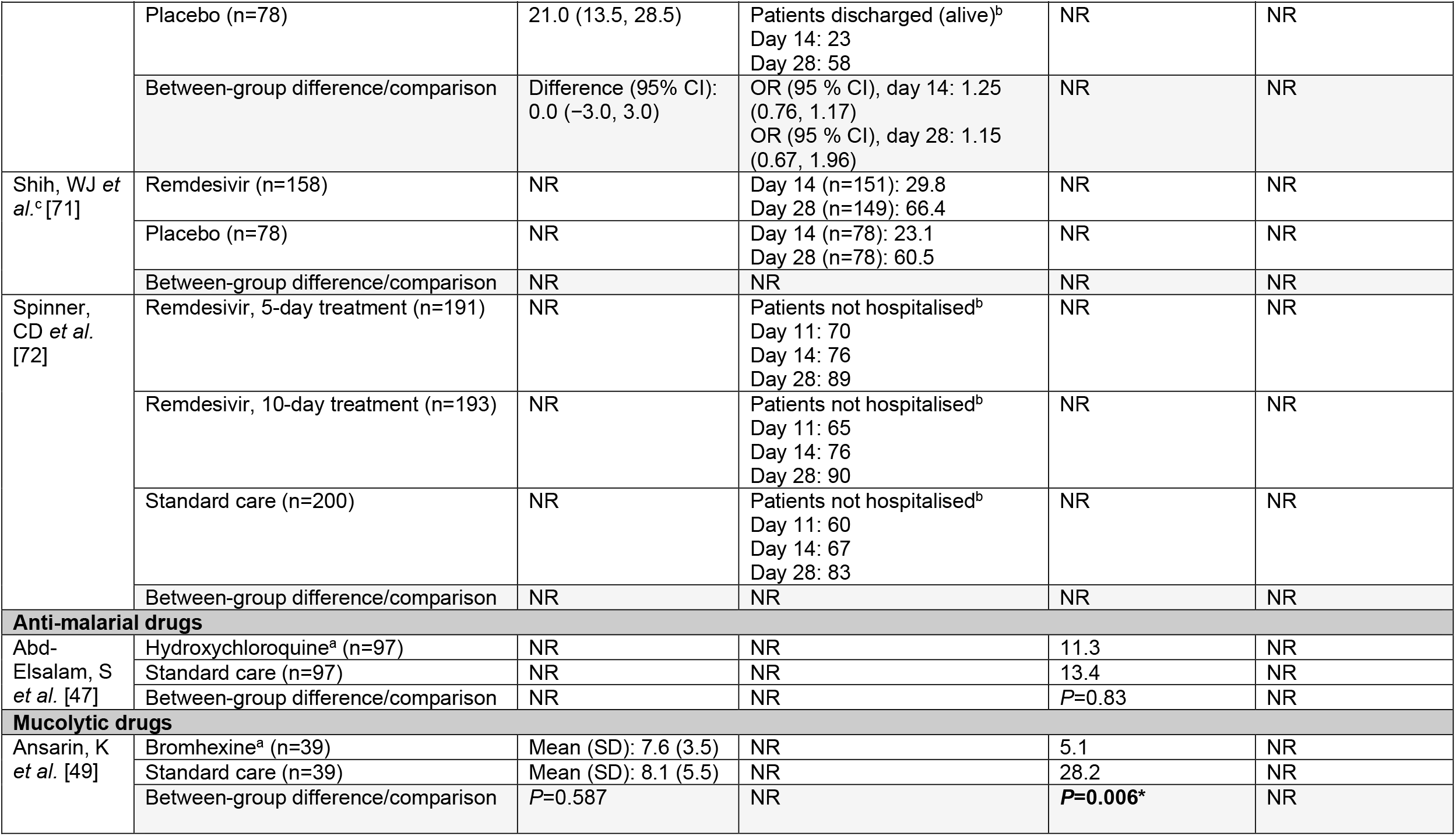
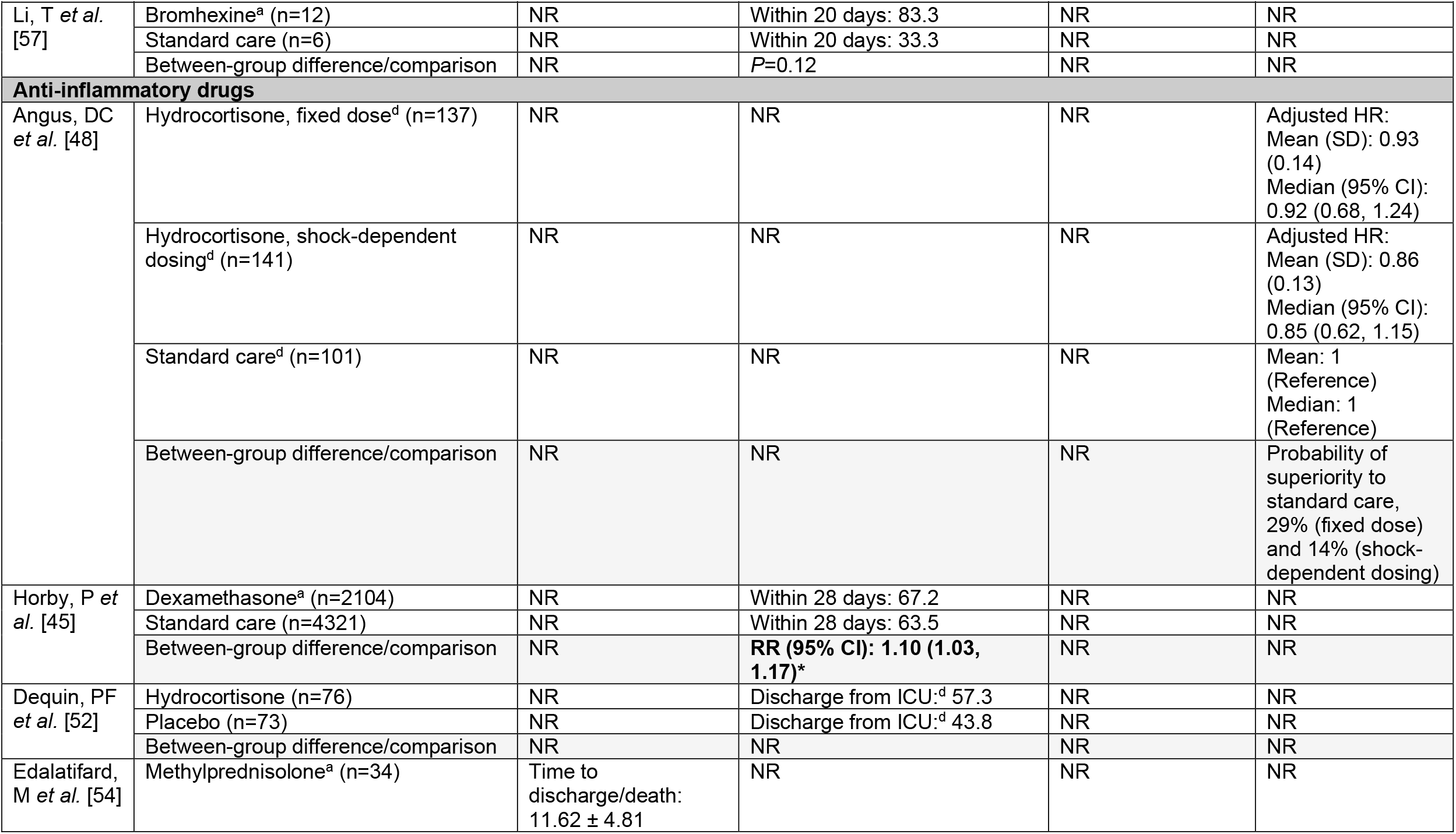
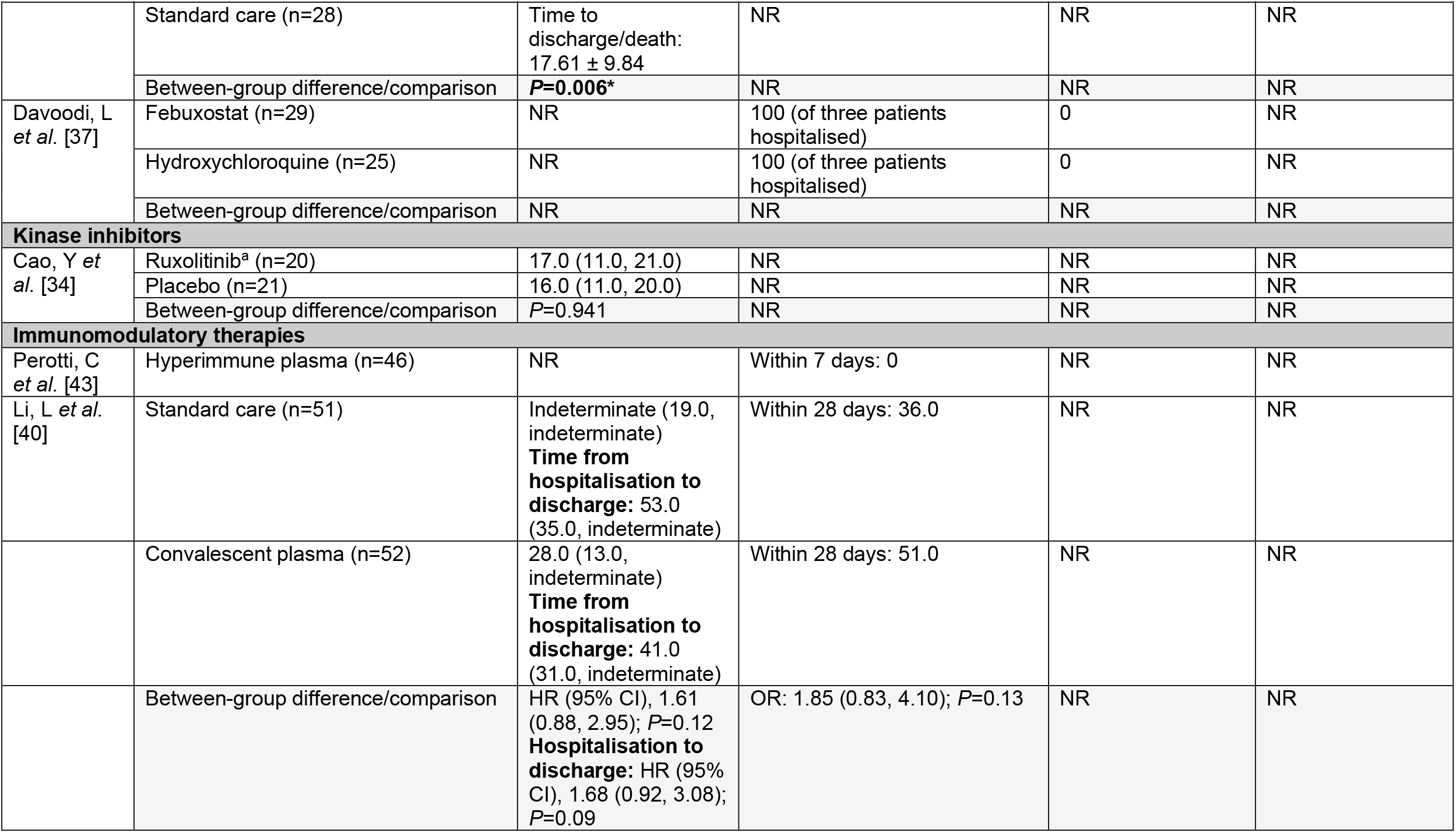
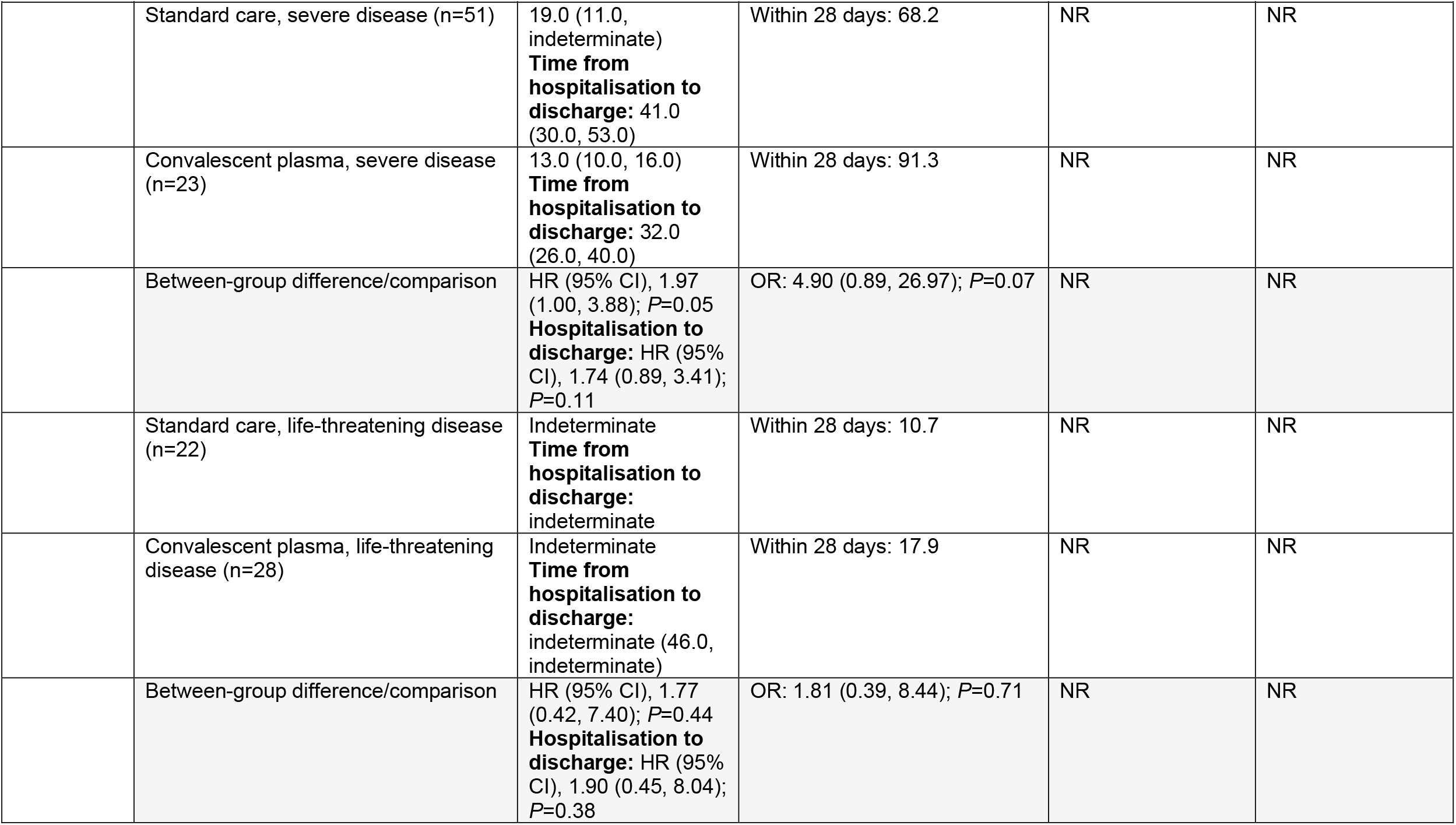
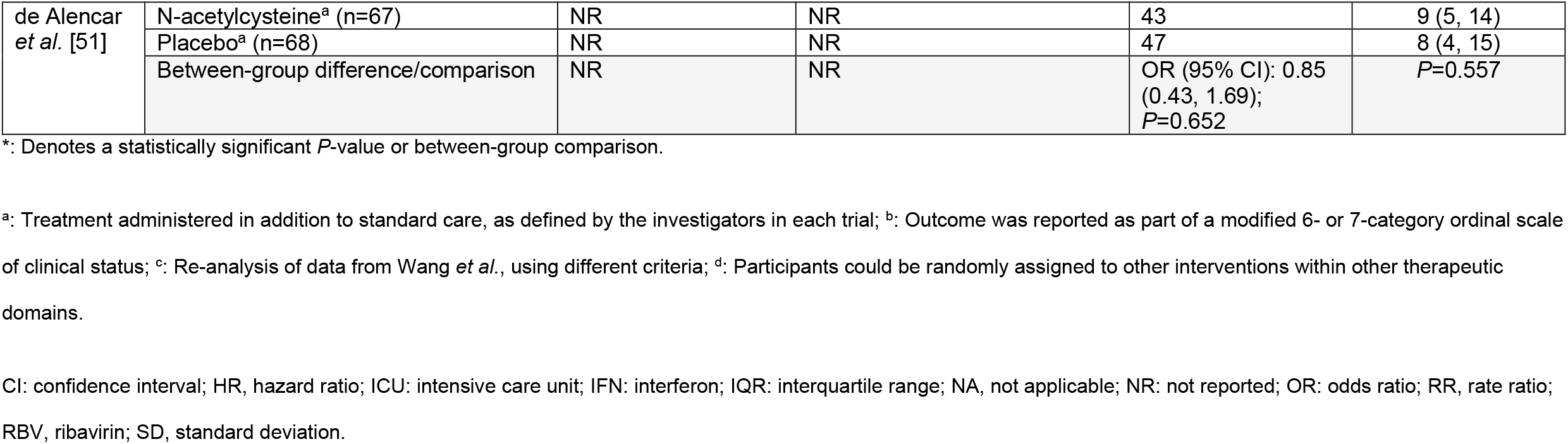
Summary of hospitalisation outcomes in trials included in the qualitative synthesis

**FIGURE 2.**
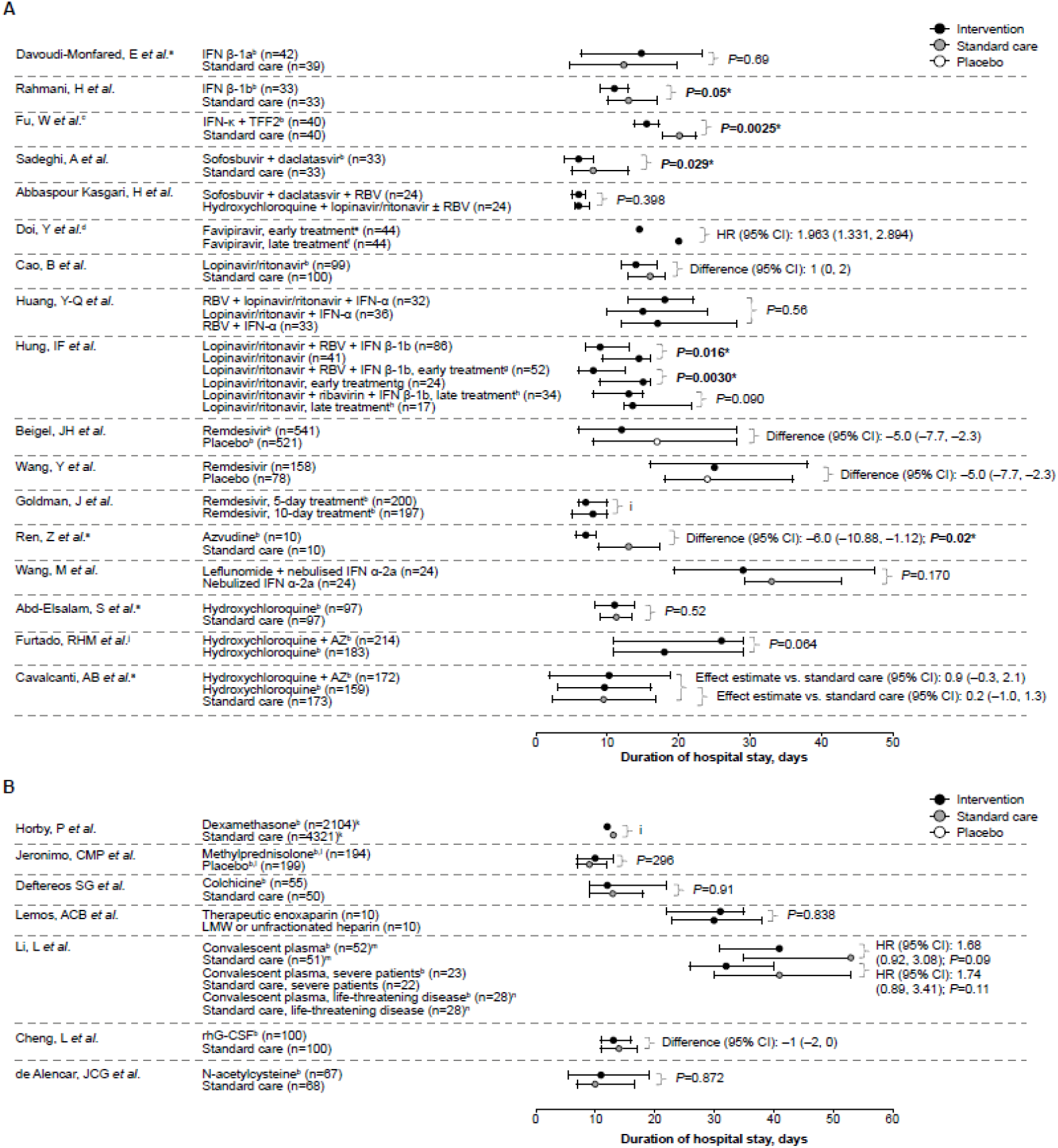
Duration of hospitalisation in trials of antivirals, anti-malarial and mucolytic drugs (A), and other therapies (B) included in the qualitative synthesis. Data are median (IQR) unless indicated otherwise. Other therapies include anti-inflammatory drugs, anticoagulants, immunomodulatory therapies and repair therapies. *: Indicates a statistically significant *P*-value. ^a^: Data are mean (SD); ^b^: Treatment administered in addition to standard care, as defined by the investigators in each trial; ^c^: Data are mean (95% CI); ^d^: *Post-hoc* analysis; ^e^: Treatment on day 1 of study participation; ^f^: Treatment on day 6 of study participation; ^g^: Treatment, <7 days from symptom onset; ^h^: Treatment ≥7 days from symptom onset; ^i^: No between-group comparison; ^j^: Outcome only assessed in survivors; ^k^: No IQR reported; ^l^: Per hospital protocol, all patients meeting ARDS criteria were given pre-emptively intravenous ceftriaxone (1 g 2x for 7 days) plus azithromycin (500 mg 1x for 5 days) or clarithromycin (500 mg 2x for 7 days), starting on day 1; ^m^: Upper IQR limit could not be determined; ^n^: Median value could not be determined, HR: 1.90 (95% CI: 0.45, 8.04); *P*=0.38. ARDS: acute respiratory distress syndrome; AZ: azithromycin; CI: confidence interval; HR: hazard ratio; IFN, interferon; IQR: interquartile range; LMW: low molecular weight; RBV: ribavirin; rhG-CSF: recombinant human granulocyte colony-stimulating factor; SD: standard deviation.

### Mortality

In total, 39 randomised and one non-randomised trial reported on mortality, either as the number of deaths that occurred during the study or as a pre-specified study endpoint, with or without a statistical comparison between groups [30-36, 38-56, 58-70, 72]. One study included [71] was a re-analysis of data from another trial (Wang *et al*. [46]) included in the qualitative synthesis. The outcomes for individual trials are shown in figure 1.

#### Antivirals

Among trials of antivirals (figure 1A), four trials (N=48–66) reported trends towards decreased mortality with interferons, sofosbuvir + daclatasvir, and triazavirin, compared with standard care or placebo in patients with COVID-19 of varying severity [61, 62, 69, 70]. Another trial (N=81) reported a significant reduction in 28-day mortality with interferon (IFN) β-1a plus standard care compared with standard care alone (19.0% *versus* 43.6%; *P*=0.015) in patients with severe COVID-19 [66]. The analysis also showed that administration of IFN β-1a early in the disease significantly reduced mortality (odds ratio [OR]: 13.5; 95% confidence interval [CI]: 1.5–118) whereas late administration did not [66].

Of four trials that assessed lopinavir/ritonavir, three trials of patients with COVID-19 of varying severity (N=86–127) reported no deaths in either treatment group [39, 41, 64]. One trial (N=199) reported a numerical but non-significant reduction in 28-day mortality with lopinavir/ritonavir *versus* standard care in patients with severe COVID-19 [33]. Of four trials that investigated remdesivir in patients with moderate or severe COVID-19, two trials (N=1062 and N=236) and the re-analysis of Wang *et al*. [71] showed no significant mortality benefit of remdesivir compared with placebo [30, 46], although one study showed a trend towards reduced mortality in patients who received treatment earlier in their disease course (within 10 days of symptom onset) [46]. The other two trials (N=397 and N=596) reported comparable mortality with 5-day and 10-day remdesivir treatment [63, 72].

#### Anti-malarial and mucolytic drugs

Among eight trials of hydroxychloroquine or its derivatives (figure 1B), one trial (N=81) reported significantly greater lethality with high doses of chloroquine diphosphate compared with low doses (log-rank: −2.183; *P*=0.03) in patients with severe COVID-19 [31]. Six trials (N=150–821) reported similar mortality with hydroxychloroquine, with or without azithromycin, compared with standard care or placebo in hospitalised [35, 47, 65] or non-hospitalised [32, 42, 44] patients with mild, mild-to-moderate or severe COVID-19. Another trial (N=447) reported no mortality benefit of adding azithromycin to hydroxychloroquine compared with hydroxychloroquine alone in patients with severe COVID-19 [55]. In a trial of the mucolytic drug, bromhexine (N=78), there was a significant reduction in mortality with bromhexine plus standard care *versus* standard care alone (0% *versus* 12.8%; *P*=0.027) in patients with COVID-19 of unspecified severity [49].

#### Anti-inflammatory drugs

Among trials of corticosteroids conducted in patients with severe COVID-19, three trials (N=149–403) showed numerical but non-significant trends towards reduced mortality with hydrocortisone or methylprednisolone compared with placebo or standard care [48, 52, 68] (figure 1C). One trial (N=62) reported significantly reduced mortality (5.9% *versus* 42.9%; *P*<0.001) with methylprednisolone plus standard care *versus* standard care alone [54] (figure 1C). In a large trial (N=6425) of patients with COVID-19 of unspecified severity, 28-day mortality was significantly decreased with dexamethasone plus standard care *versus* standard care alone, both overall (22.9% *versus* 25.7%; *P*<0.001) and in patients receiving oxygen (23.3% *versus* 26.2%) or mechanical ventilation (29.3% *versus* 41.4%) at randomisation [45] (figure 1C). The mortality benefit was greatest in patients with a longer duration of symptoms (>7 days *versus* ≤7 days; 12.3 by chi-square test for trend) [45]. Another trial (N=105), also conducted in patients with COVID-19 of unspecified severity, showed significantly increased event-free survival with the anti-inflammatory drug colchicine in combination with standard care *versus* standard care alone (97% *versus* 83% of patients after 10 days; *P*=0.03; data not shown graphically) [38].

#### Other therapies

Trials (N=20–135) investigating the kinase inhibitor, ruxolitinib [34]; the calcium release-activated calcium (CRAC) channel inhibitor, auxora [58]; the anticoagulant, enoxaparin [56]; and N-acetylcysteine, a mucolytic drug with anti-oxidant properties [51], in patients with severe COVID-19 reported no significant difference in mortality *versus* the comparator groups (figure 1C). One trial (N=200) reported a reduced 21-day mortality with recombinant human granulocyte colony-stimulating factor (rhG-CSF) added to standard care *versus* standard care alone (hazard ratio [HR]: 0.19; 95% CI: 0.04–0.88) in patients with severe COVID-19 [50].

Two studies reported on immunomodulatory therapies in patients with moderate and/or severe COVID-19 (figure 1C): one single-arm trial (N=46) reported a mortality of 6.5% with hyperimmune plasma [43], and another trial (N=103) reported numerical but non-significant trends towards decreased mortality with convalescent plasma *versus* standard care [40].

### Hospitalisation

In total, 36 randomised trials and one non-randomised trial reported on hospitalisation, either as a pre-specified study endpoint or the number of patients experiencing a particular outcome during the study, with or without a statistical comparison between groups [30, 32-35, 37-40, 42-57, 59-64, 66-70, 72]. Additionally, one study included [71] was a re-analysis of data from another trial (Wang *et al*. [46]) included in the qualitative synthesis. Outcomes reported across studies included median duration of hospitalisation (figure 2), proportion of patients discharged during the study (table 2), incidence of intensive care unit (ICU) admittance (table 2) and incidence of hospitalisation (supplementary figure 3).

**FIGURE 3.**
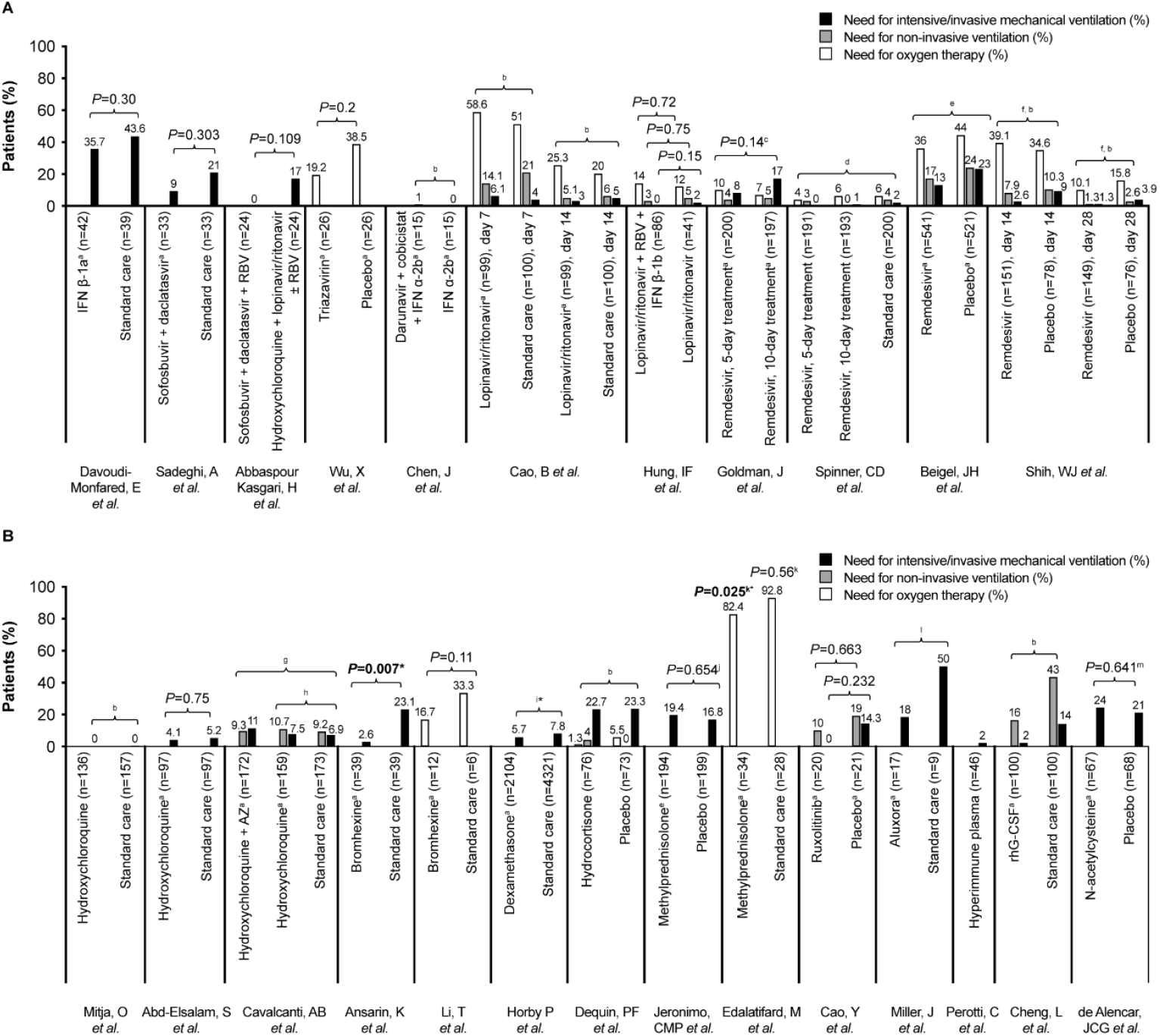
Need for ventilation in trials of antivirals (A) and other therapies (B) included in the qualitative synthesis. Other therapies include anti-malarial drugs, mucolytic drugs, anti-inflammatory drugs, anticoagulants, immunomodulatory therapies and repair therapies. Results from two studies are not presented graphically. Li Y *et al*. [41] reported need for intensive mechanical ventilation in two of 13 severe patients (15.4%); Deftereos *et al*. [38] reported need for ventilation among seven patients who met the primary clinical endpoint: in the control group, one of seven patients (14.3%) needed non-invasive mechanical ventilation and five (71.4%) were intubated and ventilated mechanically. The patient in the colchicine group who met the endpoint needed invasive mechanical ventilation. *: Denotes a significant *P*-value or other comparison. ^a^: Treatment administered in addition to standard care, as defined by the investigators in each trial; ^b^: No comparison between groups; ^c^: *P*-value for clinical status at day 14 (composite of death and ventilation requirement outcomes); ^d^: Difference in clinical status distribution *versus* standard care, odds ratio: 1.65 (95% CI: 1.09, 2.48); ^e^: Outcomes reported in patients not receiving ventilation at baseline; between-group differences (95% CI): oxygen, −8 (−24, –8); non-invasive ventilation/high-flow oxygen, −7 (−14, −1); mechanical ventilation/ECMO, −10 (−15, −4); ^f^: Re-analysis of data from Wang Y, *et al*. using different criteria; ^g^: Effect estimates *versus* standard care: need for non-invasive ventilation, 1.10 (95% CI: 0.60, 2.03); need for mechanical ventilation, 1.77 (95% CI: 0.81, 3.87); ^h^: Effect estimates *versus* standard care: need for non-invasive ventilation, 1.19 (95% CI: 0.65, 2.21); need for mechanical ventilation, 1.15 (95% CI: 0.49, 2.70); ^i^: Risk ratio: 0.77 (95% CI: 0.62, 0.95); ^j^: HR: 2.6 (95% CI: −8.6, 13.6); ^k^: Values are in comparison to baseline after 3 days after treatment. After discharge/death, the proportion of patients requiring supplementary oxygen was significantly decreased compared to baseline in both groups; ^l^: Absolute risk reduction: 32% (95% CI: −0.07, 0.71).^m^: Odds ratio (95% CI): 1.21 (0.53, 2.72) AZ: azithromycin; CI: confidence interval; ECMO: extracorporeal membrane oxygenation; HR: hazard ratio; IFN: interferon; RBV: ribavirin; rhG-CSF: recombinant human granulocyte colony-stimulating factor.

#### Antivirals

Three trials of interferons (IFN ß-1a, IFN ß-1b or IFN-κ + trefoil factor 2 [TFF2]; [N=66–81]) added to standard care in patients with moderate or severe COVID-19 reported improvements in hospitalisation outcomes, including reduced hospitalisation duration [67, 69] (figure 2A), a greater proportion of patients discharged [66, 69] (table 2) and reduced incidence of ICU admittance [66, 69] (table 2) *versus* standard care alone. Other trials (N=66 and N=88) reported reduced hospitalisation duration with sofosbuvir + daclatasvir + standard care (6 *versus* 8 days; *P*=0.029) and early *versus* late administration of favipiravir (14.5 *versus* 20 days; HR [95% CI]: 1.963 [1.331–2.894]) in patients with moderate or severe COVID-19 [70] or asymptomatic to mild COVID-19 [53], respectively (figure 2A). Patients treated with sofosbuvir + daclatasvir + standard care also had a significantly higher probability of hospital discharge by day 14 (*P*=0.041) *versus* standard care alone (table 2).

Another trial (N=127), conducted in patients with COVID-19 of unspecified severity, reported a significant reduction in median hospitalisation duration with lopinavir/ritonavir + IFN ß-1b *versus* lopinavir/ritonavir alone [39]. When patients were stratified according to the timing of treatment administration, median hospitalisation was significantly reduced in patients who received treatment within 7 days of symptom onset, but not in those who received treatment later than this [39].

Among trials of remdesivir and the re-analysis of Wang *et al*., one trial (N=1062) reported a reduced initial length of hospital stay with remdesivir *versus* placebo in patients with severe COVID-19 (median, 12 *versus* 17 days) [30] (figure 2A). There were trends towards more patients discharged with remdesivir *versus* placebo/standard care, as well as with earlier remdesivir treatment in the remaining trials [46, 63, 71, 72] (table 2), but between-group differences were either not significant or not tested. A small pilot study (N=20) also reported a reduced mean duration of hospitalisation (7 *versus* 13 days; *P*=0.02) with the antiretroviral, azvudine plus standard care *versus* standard care alone in patients with mild COVID-19 [59].

#### Anti-malarial and mucolytic drugs

Among six trials assessing hydroxychloroquine (N=194–821) [32, 35, 42, 44, 47, 55], with or without azithromycin, no benefit was seen relative to the comparator groups in terms of hospitalisation duration (figure 2A), incidence of ICU admittance (table 2) or incidence of hospitalisation (supplementary figure 3). In a trial conducted in patients with COVID-19 of unspecified severity (N=78), ICU admittance was significantly reduced with bromhexine plus standard care *versus* standard care alone (5.1% *versus* 28.2%; *P*=0.006) [49] (table 2).

#### Anti-inflammatory drugs

One trial (N=6425) reported a numerically shorter median duration of hospitalisation (12 *versus* 13 days) and a greater probability of discharge alive within 28 days with dexamethasone plus standard care *versus* standard care alone (rate ratio [95% CI]: 1.10 [1.03, 1.17]) in patients with COVID-19 of unspecified severity [45] (figure 2B; table 2). In another trial of patients with severe COVID-19 (N=403), treatment with a 7-day fixed-dose course or shock-dependent dosing of hydrocortisone were associated with reduced HRs for length of hospital and ICU stay; however, neither treatment strategy met pre-specified criteria for statistical superiority [48]. A further trial (N=62) reported a significantly reduced time to the composite outcome of hospital discharge or death with methylprednisolone plus standard care *versus* standard care alone in patients with severe COVID-19 (median, 11.6 *versus* 17.6 days; *P*=0.006) [54] (table 2). Other trials of anti-inflammatory agents (N=54–416) reported no differences in hospitalisation outcomes *versus* comparators (figure 2C; table 2; supplementary figure 3) [37, 38, 52, 68].

#### Other therapies

Among trials of other therapies, one trial (N=103) reported numerical but non-significant trends towards reduced hospitalisation duration and increased numbers of patients discharged with convalescent plasma *versus* standard care [40] (figure 2B; table 2). Trials of rhG-CSF (N=200), N-acetylcysteine (N=135), enoxaparin (N=20) and ruxolitinib (N=43) reported no significant impact of these interventions on hospitalisation outcomes *versus* standard care or placebo [34, 50, 51, 56] (figure 2B; table 2).

### Need for ventilation

In total, 30 randomised and one non-randomised trial, and one re-analysis of the study by Wang *et al*. [46], reported outcomes relating to the need for oxygen, non-invasive ventilation or intensive mechanical ventilation [30, 33-36, 38, 39, 41-43, 45-52, 54-58, 61-63, 66, 68-72]. These endpoints were reported either as pre-specified study endpoints or the number of patients receiving a particular intervention during the study, with or without a statistical comparison between groups. The outcomes for individual trials are shown in figure 3 and supplementary table 4.

#### Antivirals

Most trials of antivirals did not report a significant impact of the interventions assessed on the number of patients requiring ventilation, nor on related outcomes including duration of respiratory support (figure 3A; supplementary table 4). However, there were trends towards decreased use of ventilation with IFN therapies, sofosbuvir + daclatasvir and triazavirin (figure 3A) [61, 62, 66, 70]. Additionally, one trial (N=81) of patients with severe COVID-19 reported an increased number of patients extubated following treatment with IFN ß-1a plus standard care than with standard care alone (53.5% *versus* 11.8%; *P*=0.019) [66] (supplementary table 4). Another trial (N=1062), also conducted in patients with severe COVID-19, reported fewer patients requiring new use of oxygen (36% *versus* 44%), non-invasive ventilation (17% *versus* 24%) or intensive mechanical ventilation (13% *versus* 23%) with remdesivir *versus* placebo (figure 3A) [30].

#### Anti-malarial and mucolytic drugs

Four trials (N=194–665) reported no significant impact of hydroxychloroquine, with or without azithromycin, on reducing the need for ventilation or improving other respiratory outcomes compared with standard care, in patients with COVID-19 of a range of severities [35, 42, 47, 55] (figure 3B; supplementary table 4). Among two trials (N=78 and N=18) of bromhexine plus standard *versus* standard care alone, one reported significantly reduced numbers of patients with COVID-19 of unspecified severity requiring intensive mechanical ventilation with bromhexine (2.6% *versus* 23.1%; *P*=0.007) [49] (figure 3B). The other trial showed a numerical but non-significant trend towards reduced need for oxygen therapy with bromhexine (16.7% *versus* 33.3%; *P*=0.11) *versus* standard care, in patients with mild or moderate COVID-19 [57] (figure 3B).

#### Anti-inflammatory drugs

Among studies of anti-inflammatory drugs (figure 3B; supplementary table 4), one randomised trial (N=6425) reported a statistically significantly decreased need for invasive mechanical ventilation with dexamethasone plus standard care *versus* standard care alone in patients with COVID-19 of unspecified severity [45] (figure 3B). The risk of progression to invasive mechanical ventilation was also significantly lower with dexamethasone than with standard care (risk ratio, 0.77; 95% CI: 0.62, 0.95) [45]. Another trial (N=62) of patients with severe COVID-19 reported a significant reduction in the proportion of patients receiving oxygen after 3 days of treatment with methylprednisolone, compared with before treatment (82.4% *versus* 100%; *P*=0.025) [54] (figure 3B).

#### Other therapies

In a small trial (N=30), fewer patients with severe COVID-19 required invasive mechanical ventilation; and the composite endpoint of death or invasive mechanical ventilation occurred significantly less frequently in patients receiving auxora than in those receiving standard care (HR: 0.23; 95% CI: 0.05, 0.96; *P*<0.05) (figure 3B; supplementary table 4) [58]. In another small trial (N=20), administration of enoxaparin significantly reduced the median number of ventilator-free days compared with low molecular weight/unfractionated heparin (0 *versus* 15 days; *P*=0.028) and resulted in a higher ratio of successful liberation from mechanical ventilation after respiratory failure (HR [95% CI]: 4.0 [1.035, 15.053]; *P*=0.031) in patients with severe COVID-19 (supplementary table 4) [56]. Trials of ruxolitinib, N-acetylcysteine and rhG-CSF (N=43–200) showed no significant efficacy in reducing need for ventilation in patients with severe COVID-19 [34, 50, 51] (figure 3B; supplementary table 4).

### Quality of evidence

There was variation among trials in terms of the risk of bias in the various domains assessed (supplementary figure 4). Most trials were deemed at low risk of bias in terms of complete reporting of patient and outcomes data. However, most trials were also judged to be at high risk of bias in terms of blinding of participants and researchers to the intervention received. Only four randomised double-blind trials were considered to be at low risk of bias for all domains assessed [44, 46, 52, 68].

## Discussion

This SLR assessed evidence, up to 14 October 2020, for the efficacy of pharmacological interventions for COVID-19 in terms of mortality, hospitalisation and need for ventilation. These outcomes were selected because COVID-19 has caused significant mortality worldwide and continues to impart a substantial burden on healthcare systems [21-24, 73]. Although observational studies have yielded important and useful data, only clinical trials were included in this study to allow a synthesis of the highest quality evidence available for each of the interventions assessed.

Of 42 included trials, all but one assessed a treatment or treatment combination for patients with established COVID-19. One trial assessed the use of hydroxychloroquine as post-exposure prophylaxis in patients (N=821) with high-risk or moderate exposure to COVID-19 *versus* placebo. There was no difference in mortality or incidence of hospitalisation between groups, but the numbers of patients experiencing these events were very low (no deaths occurred and only one hospitalisation was reported in each group).

Among the trials assessing treatments in patients with COVID-19, interventions from several treatment classes showed significant effects in improving one or more of mortality, hospitalisation or need for ventilation outcomes. The most consistent effect across all outcomes assessed was reported for the corticosteroid, dexamethasone, in a preliminary report from the Randomised Evaluation of COVid-19 thERapY (RECOVERY) trial (Clinical Trials.gov: NCT04381936). The study investigators reported a significant reduction in mortality and a greater probability of discharge alive within 28 days with dexamethasone plus standard care *versus* standard care alone in patients who were receiving either invasive mechanical ventilation or oxygen at randomisation. Duration of hospitalisation was numerically shorter and the need for invasive mechanical ventilation in patients who were not already receiving this at randomisation was significantly reduced with the addition of dexamethasone to standard care [45].

Other trials of anti-inflammatory drugs included in the present review presented some evidence for efficacy in improving one or more of the outcomes assessed [38, 48, 52, 54, 68]. However, issues reported in these studies included small patient numbers [54, 68], potentially being underpowered [52], and late administration of the study treatment in some patients [68]. Further well-designed and adequately powered studies are needed to increase the body of evidence for anti-inflammatory drugs as treatment for COVID-19.

In this review, findings from trials assessing drugs acting in the early infection or early pulmonary disease phase varied depending on the type of agent evaluated, the disease stage of patients enrolled in the study and the timing of drug administration. Some efficacy in reducing mortality, hospitalisation duration and need for ventilation was seen with IFN therapies [66, 67, 69]; conversely, most trials of lopinavir/ritonavir reported no significant benefit above standard care [33, 41, 64]. Similarly, and in line with conclusions from published meta-analyses [74, 75], trials that assessed hydroxychloroquine, an antimalarial drug, or its derivatives reported no impact of these interventions on any of the outcomes of interest, and high doses of chloroquine diphosphate were associated with increased lethality compared with low doses in one trial [31].

Trials that assessed remdesivir compared with placebo or standard care also failed to show a clear benefit in terms of reducing mortality, although one trial showed a reduction in median hospitalisation duration and need for ventilation with remdesivir compared with placebo [30]. Despite the lack of strong evidence of efficacy with remdesivir, it is notable that this drug has received endorsement from regulatory bodies for treating COVID-19 on the basis of data showing improved time to recovery and fewer adverse events in remdesivir-treated patients *versus* placebo in two clinical trials included in the present review, particularly in patients with less severe disease [30, 46]. Consistent with these findings, a re-analysis of data from one trial [46] suggested that remdesivir induced good responses in patients with moderately severe, rather than critical, disease at enrolment [71]. Taken together, the data suggest that remdesivir may improve recovery time if administered early enough in the disease course; appropriate selection of patients with early-stage disease in future trials of remdesivir may help to confirm this hypothesis.

Collectively, the findings from trials of drugs acting early in the pathophysiological course of COVID-19 summarised in the present review are broadly in line with those from the SOLIDARITY trial [76]. This international, randomised trial of COVID-19 treatments concluded that all four treatments evaluated (remdesivir, hydroxychloroquine, lopinavir/ritonavir and interferons) had little or no effect on overall mortality, initiation of ventilation and hospitalisation duration in patients hospitalised with COVID-19 [76]. A notable difference is that some studies in the present review did show some efficacy of interferon therapies in reducing mortality and improving hospitalisation outcomes [66, 67, 69], suggesting that these therapies may warrant further investigation.

A key finding from this review was the relationship between the disease phase of patients with COVID-19 and the efficacy of interventions. Several studies that investigated agents targeting processes early in the disease course of COVID-19, such as viral replication, showed improved efficacy in patients who received early treatment compared with late treatment [33, 39, 46, 69]. Similarly, in a report from the RECOVERY trial, the mortality benefit of dexamethasone was greatest in patients with either the most severe disease or with the longest duration of symptoms [45], which is consistent with it acting during the inflammatory phase of the disease. These findings highlight the need for COVID-19 therapies to be tailored to patients with disease stage and severity appropriate to the mechanism of action of the intervention, as has been reported previously [77], in order for maximum efficacy to be attained.

A small number of trials included in the present review reported on other types of therapies, including kinase or CRAC channel inhibitor, anticoagulants, convalescent plasma and other immunomodulatory or repair therapies [34, 40, 43, 50, 51, 56, 58]. Some studies showed either a significant efficacy effect or a trend towards improved efficacy [34, 40, 50, 51, 56, 58]; however, in general the studies were of low quality and further clinical trials are required for a more conclusive demonstration of efficacy. In particular, although convalescent plasma has received emergency use authorisation from the Food and Drug Administration (FDA), the decision was controversial, owing to a lack of robust supporting data [12]. Only one study of convalescent plasma (N=103) was included in the current review, and did not show significant improvements in any of the outcomes assessed compared with standard care [40]. Therefore, ongoing randomised studies, including the UK’ s RECOVERY trial, will be important in providing further evidence to clarify whether convalescent plasma offers any benefit in treating COVID-19.

It was also notable that only one small clinical trial (N=20) [56] reported on an anticoagulant, enoxaparin, despite many institutions having adopted anticoagulant therapy as standard care for hospitalised patients with COVID-19. The data presented were promising and showed a reduced need for ventilation compared with patients receiving prophylactic anticoagulation. These findings support those from a recently-published large observational study, which reported an association between anticoagulant use and lower in-hospital mortality and intubation rates in patients hospitalised with COVID-19 [78]. Upcoming randomised trials will provide valuable data regarding the optimal type, duration and dose of anticoagulants for different patients.

Strengths of this SLR include the comprehensive search strategy, rigorous screening methodology and quality assessment of included studies. Although several other reviews of COVID-19 therapies have been published [7, 79, 80], we believe that our overview of the current state of the field makes an important contribution to the existing body of evidence, particularly by considering the efficacy of current treatments in the context of the disease phase in which they are used.

Limitations include the exclusion of non-English articles from searches, which may have resulted in some relevant trials not being identified. Additionally, the searches were conducted in three major databases but did not include smaller or country-specific databases. Congress abstracts were excluded, on the basis of their data being preliminary and not peer reviewed. Owing to time constraints and the fast-moving nature of the field, a review of citations in the reference lists of published SLRs and meta-analysis was not conducted; and, similarly, the review will not have captured relevant studies published after the date of the final search update.

Our ability to analyse and compare the data collected was limited by the significant variation between the trials identified, owing to differences in study design, blinding of participants, severity of illness of participants, drug doses and timing, comparators, follow-up times and outcome definitions. Most trials administered interventions in addition to standard care, which varies between countries and has changed during the pandemic as the body of evidence for the efficacy and safety of different therapies has accumulated. Similarly, the overall lack of high-quality evidence for all interventions makes it difficult to make comparisons between interventions.

Most of the trials included in this SLR were assessed as having a high risk of bias in one or more of the domains evaluated, meaning that the results of the analysis of all outcomes should be interpreted with caution. A variety of factors contributed to this: some trials were pilot [36, 57, 59, 61] or proof-of-concept [43] trials, including small patient numbers and might not have been optimally designed to show an improvement in the study outcomes. Similarly, some trials were under-powered to show significant differences between interventions [34, 40, 42, 46, 56, 64, 65]. Some trials did not include a randomised placebo control group and many trials were not double-blinded. Moreover, the follow-up period in most trials was between 14 and 28 days, and most studies did not report any longer-term data. This might have limited the ability to assess mortality and hospital discharge, particularly in patients with severe COVID-19 who often have a prolonged duration of illness.

These findings are in agreement with those reported in a living systematic review and meta-analysis of drug treatments for COVID-19, last updated in September 2020 [80]. The authors concluded that the effectiveness of most of the interventions that they assessed was uncertain, owing to the small numbers of patients enrolled in randomised controlled trials at the time and important limitations of the study designs [80]. A European Medicines Agency statement noted that small studies and compassionate programmes are unlikely to generate the required level of evidence to define the best treatment options for COVID-19 and stressed the need for multi-arm randomised controlled trials of interventions [81]. However, it should be acknowledged that the COVID-19 global public health emergency has presented an urgent need for clinical data on available interventions for the disease, with the FDA granting some substances emergency use authorisation (table 1). Thus, the available data are valuable for the purpose of practical clinical decision making.

## Conclusions

This SLR summarises evidence regarding efficacy of pharmacological interventions in terms of mortality, hospitalisation and need for ventilation in patients with COVID-19, and highlights the need for adequately powered, well-designed clinical trials to increase the quality of available evidence. The summary of findings also suggests the need to use interventions appropriate for the disease stage of COVID-19, to maximise treatment efficacy. Although no relevant data from vaccine, antibody and other novel COVID-19-specific treatments were available at the time of conducting this review, the findings from ongoing clinical trials are awaited with interest.

## Data Availability

No additional data available.

## Acknowledgements

Editorial support was provided by Rachael Cazaly, of Core Medica, London, UK, supported by AstraZeneca according to Good Publication Practice guidelines ([Link](https://www.acpjournals.org/doi/10.7326/M15-0288)).

The Sponsor was involved in the study design, analysis and interpretation of data. However, ultimate responsibility for opinions, conclusions and data interpretation lies with the authors.

