## Supplementary Materials for "Current evidence for COVID-19 therapies: a systematic literature review"

### Supplementary material

#### Supplementary File 1. Details of search terms

MEDLINE^®^ search string (searches were conducted via PubMed)

| **Search number** | **Query** | **Filters** |
| --- | --- | --- |
| 1 | Coronavirus[MeSH] OR Coronavirus[Title/Abstract] | |
| 2 | COVID*[Title/Abstract] | |
| 3 | #1 OR #2 | |
| 4 | "covid 19"[Title/Abstract] OR "Covid-19"[Title/Abstract] OR "corona virus"[Title/Abstract] OR "Novel Corona Virus"[Title/Abstract] OR "corona virus disease"[Title/Abstract] OR "Coronavirus disease"[Title/Abstract] OR "Coronavirus disease 2019"[Title/Abstract] OR nCOV[Title/Abstract] OR "n-COV"[Title/Abstract] OR "COVID19"[Title/Abstract] OR "SARS coronavirus 2"[Title/Abstract] OR "severe acute respiratory syndrome coronavirus 2"[Title/Abstract] OR "2019nCoV"[Title/Abstract] OR 2019-nCoV[Title/Abstract]OR "nCoV2019"[Title/Abstract] OR "severe acute respiratory syndrome coronavirus 2"[Title/Abstract] OR "SARS-CoV-2"[Title/Abstract] OR "SARSCoV-2"[Title/Abstract] OR "SARS-CoV2"[Title/Abstract] OR "SARSCoV19"[Title/Abstract] OR "SARS-CoV19"[Title/Abstract] OR "SARS-CoV19"[Title/Abstract] OR "HCoV-19"[Title/Abstract] OR "WN-CoV"[Title/Abstract] OR "coronavirus SARSCoV-2"[Title/Abstract] OR Coronavirus[Title/Abstract] | |
| 5 | #3 OR #4 | |
| 6 | Vaccines[MESH] | |
| 7 | vaccin*[Title/Abstract] OR prevent*[Title/Abstract] OR prophylaxis[Title/Abstract] OR prophylactic[Title/Abstract] | |
| 8 | #6 OR #7 | |
| 9 | Drug therapy[MeSH] | |
| 10 | pharmacotherap*[Title/Abstract] OR treat*[Title/Abstract] OR therap*[Title/Abstract] OR management[Title/Abstract] | |
| 11 | #9 OR #10 | |
| 12 | (drug[Title/Abstract] OR pharmacologic*[Title/Abstract] OR antiviral[Title/Abstract] OR anti-viral[Title/Abstract]) AND (treat*[Title/Abstract] OR therap*[Title/Abstract] OR agent*[Title/Abstract]) | |
| 13 | #8 OR #11 OR #12 | |
| 14 | #5 AND #13 | |
| 15 | #5 AND #13 | Other Animals |
| 16 | #14 NOT #15 | |
| 17 | 2019/11:2020/07[dp] | |
| 18 | animal OR rat* OR mouse OR guinea* OR ferret OR monkey* OR hamster* OR pig OR pigs OR rabbit OR chimpanzee* OR rodent OR mammal* OR ape OR rodent* OR mammal* OR ape OR apes | |
| 19 | #16 AND #17 | |
| 20 | human* OR patient* OR men OR women OR volunteer* | |
| 21 | #18 AND #20 | |
| 22 | #19 AND (#21 OR #20) | |
| 23 | #19 AND (#21 OR #20) | Case Reports |
| 24 | #19 AND (#21 OR #20) | Review |
| 25 | #23 OR #24 | |
| 26 | #22 NOT #25 | |
| 27 | #22 NOT #25 | English |

Embase search string

| **Search number** | **Query** |
| --- | --- |
| 1 | 'coronavirus'/exp |
| 2 | covid*:ab,ti |
| 3 | #1 OR #2 |
| 4 | 'covid 19':ab,ti OR 'covid-19':ab,ti OR 'corona virus':ab,ti OR 'novel corona virus':ab,ti OR 'corona virus disease':ab,ti OR 'coronavirus disease':ab,ti OR 'coronavirus disease 2019':ab,ti OR 'ncov':ab,ti OR 'n-cov':ab,ti OR covid19:ab,ti OR 'sars coronavirus 2':ab,ti OR '2019ncov':ab,ti OR '2019 ncov':ab,ti OR 'ncov2019':ab,ti OR 'severe acute respiratory syndrome coronavirus 2':ab,ti OR 'sars-cov-2':ab,ti OR 'sarscov-2':ab,ti OR 'sars-cov2':ab,ti OR 'sarscov19':ab,ti OR 'sars-cov19':ab,ti OR 'hcov-19':ab,ti OR 'wn-cov':ab,ti OR 'coronavirus sarscov-2':ab,ti OR coronavirus:ab,ti |
| 5 | #3 OR #4 |
| 6 | 'vaccines'/exp |
| 7 | vaccin*:ab,ti OR prevent*:ab,ti OR 'prophylaxis':ab,ti OR 'prophylactic':ab,ti |
| 8 | #6 OR #7 |
| 9 | 'drug therapy'/exp |
| 10 | pharmacotherap*:ab,ti OR treat*:ab,ti OR therap*:ab,ti OR management:ab,ti |
| 11 | #9 OR #10 |
| 12 | (drug:ab,ti OR pharmacologic*:ab,ti OR antiviral:ab,ti OR 'anti-viral':ab,ti) AND (treat*:ab,ti OR therap*:ab,ti OR agent*:ab,ti) |
| 13 | #8 OR #11 OR #12 |
| 14 | #5 AND #13 |
| 15 | #5 AND #13 AND ([animal cell]/lim OR [animal experiment]/lim OR [animal model]/lim OR [animal tissue]/lim) |
| 16 | #5 AND #13 NOT ([animal cell]/lim OR [animal experiment]/lim OR [animal model]/lim OR [animal tissue]/lim) |
| 17 | (animal:ab,ti OR rat:ab,ti OR mouse:ab,ti OR guinea*:ab,ti OR ferret:ab,ti OR monkey:ab,ti OR hamster:ab,ti OR pig:ab,ti OR rabbit:ab,ti OR chimpanzee:ab,ti OR rodent:ab,ti OR mammal*:ab,ti OR ape:ab,ti) AND (model*:ab,ti OR study:ab,ti) |
| 18 | animal*:ab,ti OR rodent*:ab,ti OR rat:ab,ti OR rats:ab,ti OR mouse:ab,ti OR mice:ab,ti OR ferret*:ab,ti OR guinea*:ab,ti OR rabbit*:ab,ti OR monkey*:ab,ti OR hamster*:ab,ti OR pig:ab,ti OR pigs:ab,ti OR chimpanze*:ab,ti OR mammal*:ab,ti OR ape:ab,ti OR apes:ab,ti |
| 19 | #17 OR #18 |
| 20 | #16 NOT #19 |
| 21 | #20 AND ([conference abstract]/lim OR [conference paper]/lim OR [conference review]/lim OR [review]/lim) AND [animals]/lim |
| 22 | #20 AND ([conference abstract]/lim OR [conference paper]/lim OR [conference review]/lim OR [review]/lim) |
| 23 | #20 NOT #22 |
| 24 | #20 NOT #22 AND [2019-2020]/py |
| 25 | #20 NOT #22 AND [2019-2020]/py AND [english]/lim |

Cochrane Library search string

| **Search number** | **Search** |
| --- | --- |
| #1 | MeSH descriptor: [Coronavirus] explode all trees |
| #2 | COVID* |
| #3 | #1 OR #2 |
| #4 | ("covid 19" OR "covid19" OR "corona virus" OR "Novel Corona Virus" OR "corona virus disease" OR "Coronavirus disease" OR "Coronavirus disease 2019" OR nCOV OR COVID19 OR "SARS coronavirus 2" OR "severe acute respiratory syndrome coronavirus 2" OR 2019nCoV OR nCoV2019 OR "severe acute respiratory syndrome coronavirus 2" OR SARSCoV19 OR "HCoV19" OR "WNCoV" OR "coronavirus SARSCoV2" OR Coronavirus):ti,ab,kw |
| #5 | #3 OR #4 |
| #6 | MeSH descriptor: [Vaccines] explode all trees |
| #7 | vaccin* OR prevent* OR "prophylaxis" OR "prophylactic" |
| #8 | #6 OR #7 |
| #9 | MeSH descriptor: [Drug Therapy] explode all trees |
| #10 | pharmacotherap* OR treat* OR therap* OR management |
| #11 | #9 OR #10 |
| #12 | ((drug OR pharmacologic* OR antiviral OR anti-viral) AND (treat* OR therap* OR agent*)):ti,ab,kw |
| #13 | #8 OR #11 OR #12 |
| #14 | #5 AND #13 |
| #15 | ((animal OR rat OR mouse OR guinea* OR ferret OR monkey OR hamster OR pig OR rabbit OR chimpanzee OR rodent OR mammal* OR ape) AND (model* OR study)):ti,ab,kw |
| #16 | (animal* OR rodent* OR rat OR rats OR mouse OR mice OR ferret* OR guinea* OR rabbit* OR monkey* OR hamster* OR pig OR pigs OR chimpanze* OR mammal* OR ape OR apes):ti,ab,kw |
| #17 | #15 OR #16 |
| #18 | #14 NOT #17 with Cochrane Library publication date Between Oct 2019 and Dec 2020 |

PubMed Central search string

| **Search number** | **Query** |
| --- | --- |
| #1 | Search ("covid 19"[Title/Abstract] OR "covid 19"[Title/Abstract] OR "corona"[Title/Abstract] OR "coronae"[Title/Abstract] OR "coronas"[Title/Abstract] OR "corona virus"[Title/Abstract] OR "Novel Corona Virus"[Title/Abstract] OR "corona virus disease"[Title/Abstract] OR "SARS-CoV-2"[Title/Abstract] OR "Coronavirus disease"[Title/Abstract] OR "Coronavirus disease 2019"[Title/Abstract] OR "nCOV"[Title/Abstract] OR "n-COV"[Title/Abstract]) |
| #2 | Search Vaccines[MeSH] |
| #3 | Search (vaccin*[Title/Abstract] OR prevent*[Title/Abstract] OR "prophylaxis"[Title/Abstract] OR "prophylactic"[Title/Abstract]) |
| #4 | Search (#2 OR #3) |
| #5 | Search Drug therapy[MeSH] |
| #6 | Search (pharmacotherap*[Title/Abstract] OR treat*[Title/Abstract] OR therap*[Title/Abstract] OR management[Title/Abstract]) |
| #7 | Search (#5 OR #6) |
| #8 | Search ((drug[Title/Abstract] OR pharmacologic*[Title/Abstract] OR antiviral[Title/Abstract] OR anti-viral[Title/Abstract]) AND (treat*[Title/Abstract] OR therap*[Title/Abstract] OR agent*[Title/Abstract])) |
| #9 | Search (#4 OR #7 OR #8) |
| #10 | Search (#1 AND #9) |
| #11 | Search ((animal OR rat OR mouse OR guinea* OR ferret OR monkey OR hamster OR pig OR rabbit OR chimpanzee OR rodent OR mammal* OR ape) AND (model* OR study)) |
| #12 | Search (animal* OR rodent* OR rat OR rats OR mouse OR mice OR ferret* OR guinea* OR rabbit* OR monkey* OR hamster* OR pig OR pigs OR chimpanze* OR mammal* OR ape OR apes) |
| #13 | Search (#11 OR #12) |
| #14 | Search (#10 NOT #13) |
| #15 | Search (case* AND (report* OR stud* OR histor* OR observation* OR series)) |
| #16 | Search (#14 NOT #15) |
| #17 | Search (#14 NOT #15) Filters: Publication date from 2020/07/01 to 2020/10/05 |

#### Supplementary Table 1. Details of interventions and categorisation

| **Disease stage** | **Interventions** |
| --- | --- |
| **Early infection/early pulmonary phase** | **Antivirals:** non-specific (interferons); broad-spectrum (favipiravir, ribavirin, sofosbuvir + daclatasvir, triazavirin); antiretrovirals (lopinavir/ritonavir, remdesivir, darunavir/cobicistat, azvudine); antiviral combinations (lopinavir/ritonavir + interferons); other (leflunomide)^a^ |
|  | **Anti-malarial drugs:** hydroxychloroquine and derivatives |
|  | **Mucolytic drugs:** bromhexine |
| **Late pulmonary phase/host inflammatory response phase/repair phase** | **Anti-inflammatory drugs:** non-specific (dexamethasone, hydrocortisone, methylprednisolone); other (febuxostat, colchicine) |
|  | **Kinase inhibitors:** ruxolitinib |
|  | **Calcium release-activated calcium channel inhibitors:** auxora |
|  | **Anticoagulants:** enoxaparin |
|  | **Immunomodulatory therapies:** convalescent plasma, hyperimmune plasma |
|  | **Repair therapies:** N-acetylcysteine |

^a^Leflunomide is an immunosuppressant that may also exhibit antiviral activity.

#### Supplementary Table 2. Characteristics of studies in articles not included in the qualitative synthesis

| **Study** | **Study design** | **No. of participants** | **Country** | **Mean age, years** | **Male, %** | **Patients + disease state** | **Intervention(s) (mode of administration)** | **Comparator (mode of administration)** | **Reason for exclusion** |
| --- | --- | --- | --- | --- | --- | --- | --- | --- | --- |
| Zhu, FC *et al.* [1, 2] | Non-randomised, open label phase I dose escalation trial | 108 | China | Low-dose, 37.2; middle-dose, 36.3; high-dose, 35.5 | 51.0 | Healthy adults, no previous SARS-CoV-2 infection | Non-replicating adenovirus type-5 (Ad5) vectored COVID-19 vaccine, intramuscular injection | None | No outcomes of interest reported |
| Zhu, FC *et al.* [1, 2] | Randomised, double-blind, placebo-controlled phase II trial | 508 | China | 39.7 | 50.0 | Healthy adults, no previous SARS-CoV-2 infection | Non-replicating adenovirus type-5 (Ad5) vectored COVID-19 vaccine, intramuscular injection | Placebo (containing only vaccine excipients), intramuscular injection | No outcomes of interest reported |
| Humeniuk, R *et al.* [3] | Randomised, blinded, placebo-controlled (single- and multiple-dose) phase I trials | Single dose: 96 Multiple dose: 24 | US | Single dose: 44.0 Multiple dose: 44.0 | Single dose: 58.3 Multiple dose: 58.3 | Healthy adults | Remdesivir (IV) | Placebo (IV) | No outcomes of interest reported |
| Cai, Q *et al.* [4] | Non-randomised open-label trial; phase not reported | 80 | China | Median, 47.0 | 43.8 | Moderate disease; hospitalised | Favipiravir (oral). In addition, all participants received IFN-α1b (aerosol inhalation) | Lopinavir/ritonavir (oral). In addition, all participants received IFN-α1b (aerosol inhalation) | No outcomes of interest reported |
| Gautret, P *et al.* [5] | Non-randomised open-label phase III trial | 36 | France | 45.1 | 41.7 | Unspecified disease severity; hospitalised | Hydroxychloroquine with/without azithromycin (oral) | Standard care | No outcomes of interest reported |
| Antorini, S *et al.* [6] | Non-randomised, single-arm, open-label trial | 35 | Italy | Median, 63.0 | 74.3 | Severe disease; hospitalised | Remdesivir (IV). Patients could continue their existing treatments, apart from lopinavir/ritonavir which was required to be discontinued | None | Randomised trial data available for interventions reported |
| Erkurt, MA *et al.* [7] | Non-randomised, single-arm trial | 26 | Turkey | 67.4 | 69.2 | Severe disease; patients in ICU | Convalescent plasma therapy (IV) | None | Randomised trial data available for interventions reported |
| Abolghasemi, H *et al.* [8] | Non-randomised, open-label trial | 189 | Iran | Plasma, 54.4; control, 56.8 | Plasma, 58.3; control, 50.0 | Unspecified disease severity; hospitalised patients | Convalescent plasma therapy (IV) in addition to standard care (including lopinavir/ritonavir, hydroxychloroquine and an anti-inflammatory agent) | Standard care | Randomised trial data available for interventions reported |
| Liu, X *et al.* [9] | Randomised, open label phase IV trial | 31 | China | 56.0 | 67.7 | Non-severe to critical COVID-19; hospitalised | Dipyridamole (oral) in addition to standard care | Standard care | Unclear reporting of the study design, including blinding |
| Leng, Z *et al.* [10] | Randomised, open label (pilot) phase I/II trial | 10 | China | 59.4 | 40.0 | Mild, moderate and severe disease; hospitalised | MSC treatment (IV) | Placebo (IV) | Very low patient number (N=10) Unclear reporting with most outcomes data reported for one critically severe patient |
| Abella, BS *et al.* [11] | Randomised, double-blind, placebo-controlled phase II study | 132 | US | Median, 33; | 31.0 | Mostly healthy/asymptomatic healthcare workers in hospital settings | Hydroxychloroquine (oral) | Placebo (oral) | No outcomes of interest reported |
| Anderson, EJ *et al.* [12] | Non-randomised, open label, phase I trial | 40 | US | 68.7 | 48.0 | Healthy older patients (≥56 years) | mRNA-1273 (IM injection) | None | No outcomes of interest reported |
| Bradfute, SB *et al.* [13] | Single-arm, open label phase II trial | 12 | US | Median, 52 | 66.7 | SARS-CoV-2 infected patients with respiratory symptoms and in need of supplemental oxygen; hospitalised | Convalescent plasma therapy (IV) | None | No outcomes of interest reported |
| Keech, C *et al.* [14] | Randomised, blinded, placebo-controlled phase I/II trial | 131 Vaccine + adjuvant: 83 Vaccine without adjuvant: 25 Placebo: 23 | Australia | 30.8 | 50.4 | Healthy adults (18–59 years), no previous SARS-CoV-2 infection | NVX-CoV2373 nanoparticle vaccine (IM injection) | Placebo | No outcomes of interest reported |
| Logunov, DY *et al.* [15] | Non-randomised, open label phase I/II trial | 76 | Russia | Gam-COVID-Vac – rAd26-S, 27.8; rAd5-s, 25.3; rAd26-S+rAd5-S, 26.4  Gam-COVID-Vac-Lyo – rAd26-S, 31.4; rAd5-S, 27.0; rAd26-s+rAd5-S, 26.7 | Gam-COVID-Vac – rAd26-S, 100; rAd5-S, 100; rAd26-S+rAd5-S, 70  Gam-COVID-Vac-Lyo – rAd26-S, 56; rAd5-S, 22; rAd26-s+rAd5-S, 70 | Healthy adults (18–60 years), no previous SARS-CoV-2 infection | rAd26-S and rAd5-S (IM injection) | None | No outcomes of interest reported |
| Xia, S *et al.* [16] | Randomised, double-blind, placebo-controlled, phase I/II trial | Phase I: 96 Phase II: 224 | China | Phase I, 41.2; Phase II, 43.5 Overall: 42.8 | Phase I, 39.6; Phase II, 36.6 Overall: 37.5 | Healthy adults (18–59 years), no previous SARS-CoV-2 infection | Inactivated whole-virus COVID-19 vaccine (IM injection) | Placebo (only used in phase II study) | No outcomes of interest reported |
| Zheng, F *et al.* [17] | Randomised, open label, parallel-group phase IV trial | 89 | China | Novaferon, 46.5; lopinavir/ritonavir + novaferon, 50.0; Lopinavir/ritonavir, 37.0 | Novaferon, 56.7, lopinavir/ritonavir + novaferon, 43.3; Lopinavir/ritonavir, 41.4 | Hospitalised adult patients with moderate-to-severe SARS-CoV-2 infection | Novaferon (aerosolized inhalation) Novaferon aerosolized inhalation + lopinavir/ritonavir (oral) | Lopinavir/ritonavir (oral) | No outcomes of interest reported |
| Fu, W [18] | Non-randomised, open label pilot trial; phase not specified | 33 | China | Median, 50 | 57.6 | Hospitalised patients with moderate SARS-CoV-2 infection | IFN-κ + TFF2 (aerosol inhalation) | Standard care | Non-randomised study |

COVID-19: Coronavirus disease 2019; ICU: intensive care unit; IFN, interferon; IM, intramuscular; IV: intravenous; mRNA: messenger RNA; MSC, mesenchymal stem cells; SARS-CoV-2: severe acute respiratory syndrome coronavirus 2; TFF2: trefoil factor 2.

#### Supplementary Table 3. Characteristics of studies in articles included in the qualitative synthesis

| **Study** | **Study design** | **No. of participants** | **Country** | **Mean age, years** | **Male, %** | **Patients + disease state** | **Intervention(s) (mode of administration)** | **Comparator (mode of administration)** |
| --- | --- | --- | --- | --- | --- | --- | --- | --- |
| **Antivirals** | | | | | | | | |
| Hung IF *et al.* [19] | Randomised, open label phase II/IIb study | 127 | Hong Kong | Median, 52.0 | 54.0 | Unspecified disease severity; hospitalised | Lopinavir/ritonavir (NG tube) + RBV (inhaled) + IFN beta-1b (SC) | Lopinavir/ritonavir (NG tube) |
| Goldman, J *et al.* [20] | Randomised, open label phase III study | 397 | Multinational | Median, 5-day group: 61.0 10-day group: 62.0 | 5-day group: 60.0 10-day group: 68.0 | Severe disease, evidence of pneumonia; hospitalised | Remdesivir (IV) in addition to supportive therapy | No comparator (study compared 5 or 10 days of remdesivir treatment) |
| Beigel, JH *et al.* [21] | Randomised, double-blind, placebo-controlled phase III study | 1062 | Multinational | 58.9 | 64.4 | Severe disease; hospitalised | Remdesivir (IV) in addition to supportive care (including other treatments for COVID-19) | Placebo in addition to supportive care (including other treatments for COVID-19) |
| Wang, Y *et al.* [22] | Randomised, double-blind, placebo-controlled phase III study | 236 | China | Median, 65.0 | Remdesivir: 56.0 Placebo: 65.0 | Severe COVID-19 pneumonia; hospitalised | Remdesivir (IV) | Placebo |
| Chen, J *et al*. [23] | Randomised, parallel assignment, open label phase III study | 30 | China | 47.2 | 60.0 | Mild COVID-19 pneumonia; hospitalised | Darunavir (oral) + cobicistat (oral) + interferon alpha 2b (inhaled) in addition to standard care | Interferon alpha 2b (inhaled) in addition to standard care |
| Li, Y *et al.* [9] | Randomised, partially blinded phase IV study | 86 | China | 49.4 | 46.5 | Mild/moderate disease; hospitalised | Lopinavir/ritonavir (oral) or arbidol (oral) in addition to standard care | Standard care |
| Cao, B *et al.* [24] | Randomised, open label study; phase not reported | 199 | China | Median, 58.0 | 60.3 | Severe disease; hospitalised | Lopinavir/ritonavir (oral) in addition to standard care | Standard care |
| Huang, Y-Q *et al.* [25] | Randomised, open label study; phase not reported | 101 | China | 42.5 | 46.0 | Mild to moderate disease; hospitalised | RBV (initial dose IV and then oral) + lopinavir/ritonavir (oral) + IFN-a (inhaled) | RBV (IV) + IFN-a (inhaled)  Lopinavir/ritonavir (oral) + IFN-a (inhaled) |
| Spinner, CD *et al.* [26] | Randomised, open label phase III study | 596 | USA | Median: 5-day remdesivir, 58; 10-day remdesivir, 56; standard care, 57 | 5-day remdesivir, 60; 10-day remdesivir, 61; standard care, 63 | Moderate disease; hospitalised | Remdesivir (IV) | Standard care |
| Shih, WJ *et al.*^a^ [27] | Randomised, double-blind, placebo-controlled phase III study | 231 patients included in re-analysis of data from Wang, Y *et al.* [22] | China | NR | NR | Severe Covid-19 pneumonia; hospitalised | Remdesivir (IV) | Placebo |
| Abbaspour Kasgari, H *et al.* [28] | Randomised, open label phase III study | 48 | Iran | Median: sofosbuvir/ daclatasvir, 45; control, 60 | Sofosbuvir/ daclatasvir, 46; control, 29 | Moderate disease; hospitalised | Sofosbuvir + daclatasvir + ribavirin (oral) | Standard care |
| Davoudi-Monfared, E *et al.* [29] | Randomised, open label phase III study | 81 | Iran | IFN-β, 56.0; control, 59.5 | 54.3 | Severe disease; hospitalised | Interferon β-1a (SC injections) in addition to standard care | Standard care |
| Sadeghi, A *et al.* [30] | Randomised, open label phase III study | 66 | Iran | Median, 58 | 52 | Moderate/severe disease; hospitalised | Sofosbuvir + daclatasvir (oral) in addition to standard care | Standard care |
| Wu, X *et al.* [31] | Randomised, double-blind phase III study | 52 | China | Median, 58 | 50 | Unspecified disease severity; hospitalised | Triazavirin (oral) in addition to standard care | Placebo in addition to standard care |
| Wang, M *et al.* [32] | Randomised, open label phase III study | 48 | China | Median: combination group, 56.0; control group, 55.5 | Combination group, 54.2; control group, 37.5 | Mild to severe disease; hospitalised | Leflunomide (oral) plus nebulized IFN alpha-2a | Nebulized IFN alpha-2a (inhalation) |
| Doi, Y *et al.* [33] | Randomised, open label study | 88 | Japan | Median, 50.0 | 61.4 | Asymptomatic or mild disease; hospitalised | Favipiravir (oral) | No comparator |
| Rahmani, H *et al.* [34] | Randomised, open label study | 66 | Iran | Median, 60 | 59.1 | Severe disease; hospitalised | IFN β-1b (SC injections) in addition to standard care | Standard care: lopinavir/ritonavir or atazanavir/ritonavir plus hydroxychloroquine (oral) |
| Fu, W *et al.* [35] | Randomised, open label study | 80 | China | 35.3 | 63.8 | Moderate disease; hospitalised | IFN-k and TFF2 (aerosol inhalation) in addition to standard care | Standard care |
| Ren, Z *et al.* [36] | Randomised, open label study | 20 | China | Median, 52 | 60 | Mild disease; hospitalised | Azvudine (oral) plus symptomatic treatment | Standard antiviral treatment plus symptomatic treatment |
| **Antimalarial drugs** | | | | | | | | |
| Borba, M *et al. [37]* | Randomised, parallel assignment, double-blind phase II/IIb study | 81 | Brazil | 51.1 | 75.3 | Severe disease; hospitalised | Chloroquine diphosphate (oral/NG tube) in addition to standard care | No comparator |
| Boulware, DR *et al.* [38] | Randomised, double-blind, placebo-controlled phase III study | 821 | US/Canada | Median, 40.0 | 48.4 | Asymptomatic adults with occupational/household exposure to COVID-19 | Hydroxychloroquine (oral) | Placebo |
| Skipper, C *et al.* [39] | Randomised, double-blind, placebo-controlled phase III study | 491 | North America | Median, 40.0 | 44.0 | Mild COVID-19; non-hospitalised | Hydroxychloroquine (oral) | Placebo |
| Mitja, O *et al.* [40] | Randomised, open label phase III study | 293 | Spain | 41.6 | 31.4 | Mild COVID-19; outpatients | Hydroxychloroquine (oral) | Standard care |
| Cavalcanti *et al.* [41] | Randomised, open label phase III study | 665 | Brazil | 50.3 | 58.3 | Mild-to-moderate COVID-19; hospitalised | Hydroxychloroquine (oral) in addition to standard care  Hydroxychloroquine (oral) + azithromycin (oral) in addition to standard care | Standard care |
| Tang, W *et al.* [42] | Randomised, open label phase IV study | 150 | China | 46.1 | 54.7 | Mild, moderate and severe disease; hospitalised | Hydroxychloroquine (oral) in addition to standard care | Standard care |
| Abd-Elsalem, S *et al.* [43] | Randomised, open label, phase III study | 194 | Egypt | 40.7 | 58.8 | Mild, moderate and severe disease; hospitalised | Hydroxychloroquine (oral) in addition to standard care | Standard care |
| Furtado, RHM *et al.* [44] | Randomised, open label phase III study | 447 | Brazil | Median, 59.8 | 66 | Severe disease; hospitalised | Azithromycin + hydroxychloroquine (oral) in addition to standard care | Hydroxychloroquine in addition to standard care |
| **Mucolytic drugs** | | | | | | | | |
| Li, T *et al.* [45] | Randomised, open label study; phase not reported | 18 | China | Median, 52 | 77.8 | Mild or moderate disease; hospitalised | Bromhexine hydrochloride (oral) in addition to standard care | Standard care |
| Ansarin, K *et al.* [46] | Randomised, open label phase III study | 78 | Iran | Treatment group, 58.4; standard group, 61.1 | 54.4 | Unspecified disease severity; hospitalised | Bromhexine hydrochloride (oral) in addition to standard care | Standard care |
| **Anti-inflammatory drugs** | | | | | | | | |
| Horby, P *et al.* [47] | Randomised, open label phase III study | 6425 | UK | 66.1 | 64.0 | Unspecified disease severity; hospitalised | Dexamethasone (IV and oral) in addition to standard care | Standard care |
| Deftereos SG *et al.* [48] | Randomised, open label phase II/IIb study | 105 | Greece | Median, 64.0 | 58.1 | Unspecified disease severity; hospitalised | Colchicine (oral) in addition to standard care | Standard care |
| Davoodi, L *et al.* [49] | Randomised, double-blind phase III study | 54 | Iran | 57.7 | 59.3 | Mild to moderate COVID-19; outpatients | Febuxostat (oral) in addition to supportive care (acetaminophen 325 mg, as needed, for controlling fever) | Hydroxychloroquine in addition to supportive care (acetaminophen 325 mg, as needed, for controlling fever) |
| Edalatifard, M *et al.* [50] | Randomised, single blind phase II study | 62 | Iran | 58.5 | 62.9 | Severe disease; hospitalised | Methylprednisolone (IV) in addition to standard care | Standard care |
| Dequin, PF *et al.* [51] | Randomised, double blind phase III study | 149 | France | 62.2 | 69.8 | Severe disease; hospitalised | Hydrocortisone (IV) (adjunctive treatments were also permitted) | Placebo (adjunctive treatments were also permitted) |
| Jeronimo, CMP *et al.* [52] | Randomised, double-blind phase IIb study | 416 | Brazil | 55 | 64.6 | Severe disease; hospitalised | Methylprednisolone (IV) in addition to standard care | Placebo in addition to standard care |
| Angus, DC *et al.* [53] | Randomised, open label phase IV study | 403 | Multi-national | 60 | 71 | Severe disease; hospitalised | Hydrocortisone (IV) (participants could be randomly assigned to other interventions within other therapeutic domains) | Standard care (participants could be randomly assigned to other interventions within other therapeutic domains) |
| **Kinase inhibitors** | | | | | | | | |
| Cao, Y *et al.* [54] | Randomised, single-blind, placebo-controlled phase II/IIb study | 43 | China | 63 | 58.5 | Severe disease; hospitalised | Ruxolitinib (oral) in addition to standard care | Placebo in addition to standard care |
| **Calcium release-activated calcium channel inhibitors** | | | | | | | | |
| Miller, J *et al.* [55] | Randomised, open label phase II study | 30 | US | Arm A – Auxora, 59; standard care, 61  Arm B – Auxora, 64; standard care, 36 | Arm A – Auxora, 41; SOC, 56  Arm B – Auxora, 33; SOC, 100 | Severe disease; hospitalised | Auxora (IV) in addition to standard care | Standard care |
| **Anticoagulants** | | | | | | | | |
| Lemos, ACB *et al.* [56] | Randomised, open label phase II study | 20 | Brazil | Standard prophylactic anticoagulation, 58; therapeutic enoxaparin, 55 | Standard prophylactic anticoagulation, 70; therapeutic enoxaparin, 90 | Severe disease; hospitalised | Enoxaparin (SC) | Standard anticoagulant thromboprophylaxis |
| **Immunomodulatory therapies** | | | | | | | | |
| Li, L *et al.* [57] | Randomised, open label; phase not reported | 103 | China | Median, 70 | 58.3 | Severe disease; hospitalised | Convalescent plasma (IV) and supportive treatments | Standard care |
| Perotti, C *et al.* [58] | Single-arm proof of concept study; phase not reported | 46 | Italy | 63.0 | 61.0 | Moderate to severe disease; hospitalised | Hyperimmune plasma (IV) | None |
| **Repair therapies** | | | | | | | | |
| Cheng, L *et al.* [59] | Randomised, open label study | 200 | China | Median, 45 | 56 | Severe disease; hospitalised | rhG-CSF (SC injection) in addition to standard care | Standard care |
| De Alencar, JCG *et al.* [60] | Randomised, double-blind study | 135 | Brazil | Median: N-acetylcysteine, 59; placebo, 58 | N-acetylcysteine, 67; placebo, 54 | Severe disease; hospitalised | N-acetylcysteine (IV) in addition to standard care | Placebo in addition to standard care |

^a^: Re-analysis of the clinical trial reported by Wang *et al.,* using different criteria.
COVID-19: Coronavirus disease 2019; IFN, interferon; IV: intravenous; NG: nasogastric; NR: not reported; RBV: ribavirin; rhG-CSF: recombinant human granulocyte colony-stimulating factor; SC: subcutaneous; SOC: standard of care; TFF2: trefoil factor 2.

#### Supplementary Table 4. Need for ventilation: other outcomes reported in trials included in the qualitative synthesis

| **Study** | **Interventions** | **Median (IQR) duration of respiratory support, days** | **No. days free from respiratory report, days** | **Other outcomes** |
| --- | --- | --- | --- | --- |
| **Antivirals** | | | | |
| Davoudi-Monfared, E *et al.* [29] | IFN ß-1a^a^ (n=42) | Mean (SD), IMV: 10.8 (5.38) | NR | Extubation rate, % of intubated patients: 53.5 |
|  | Standard care (n=39) | Mean (SD), IMV: 7.82 (7.84) | NR | Extubation rate, % of intubated patients: 11.8 |
|  | Between-group difference/comparison | *P*=0.47 | NR | ***P*=0.019*** |
| Rahmani, H *et al.* [34] | IFN β-1b^a^ (n=33) | NR | NR | Intubation requirement, % of patients: 6.1 |
|  | Standard care (n=33) | NR | NR | Intubation requirement, % of patients: 18.2 |
|  | Between-group difference/comparison | NR | NR | *P*=0.12 |
| Abbaspour Kasgari, H *et al.* [28] | Sofosbuvir + daclatasvir + RBV (n=24) | IMV: 0 | NR | NR |
|  | Hydroxychloroquine + lopinavir/ritonavir +/- RBV (n=24) | IMV: 2.5 (1.5, 7.0) | NR | NR |
|  | Between-group difference/comparison | NR | NR | NR |
| Cao, B *et al.* [24] | Lopinavir/ritonavir^a^ (n=99) | Oxygen: 12.0 (9.0, 16.0) IMV: 4.0 (3.0, 7.0) | NR | NR |
|  | Standard care (n=100) | Oxygen: 13.0 (6.0, 16.0) IMV: 5.0 (3.0, 9.0) | NR | NR |
|  | Between-group difference/comparison | Difference (95% CI) Oxygen: 0 (−2.0, 2.0) IMV: −1.0 (−4.0, 2.0) | NR | NR |
| Wang, Y *et al.* [22] | Remdesivir (n=158) | Oxygen: 19 (11, 30) IMV: 7 (4, 16) Ventilation (survivors): 19.0 (5.0, 42.0) Ventilation (non-survivors): 7.0 (2.0, 11.0) | NR | NR |
|  | Placebo (n=78) | Oxygen: 21 (14, 30.5) IMV: 15.5 (6.0, 21.0) Ventilation (survivors): 42.0 (17.0, 46.0) Ventilation (non-survivors): 8.0 (5.0, 16.0) | NR | NR |
|  | Between-group difference/comparison | Difference (95% CI) Oxygen: −2.0 (−6.0, 1.0)  IMV: −4.0 (−14.0, 2.0)  Ventilation (survivors): −12.0 (−41.0, 25.0)  Ventilation (non-survivors): −2.5 (−11.0, 3.0) | NR | NR |
| Beigel, JH *et al.* [21] | Remdesivir^a^ (n=541) | Oxygen: 4 (2, 12) Non-invasive ventilation/high-flow oxygen: 3 (1, 10.5) IMV/ECMO: 21.5 (9, 28) | NR | NR |
|  | Placebo (n=521) | Oxygen: 5.5 (1, 15) Non-invasive ventilation/high-flow oxygen: 4 (2, 23.5) IMV/ECMO: 23 (12, 28) | NR | NR |
|  | Between-group difference/comparison, | Difference (95% CI) Oxygen: −1.0 (−7.6, 5.6) Non-invasive ventilation/high-flow oxygen: −1.0 (−4.0, 2.0) IMV: 1.0 (−6.0, 8.0) | NR | NR |
| **Anti-malarial drugs** | | | | |
| Calvacanti *et al.* [41] | Hydroxychloroquine + AZ^a^ (n=169) | NR | 11.1 (4.9) | NR |
|  | Hydroxychloroquine^a^ (n=157) | NR | 11.2 (4.9) | NR |
|  | Standard care (n=171) | NR | 11.1 (4.9) | NR |
|  | Between-group difference/comparison | NR | Effect estimate (95% CI), hydroxychloroquine + AZ *versus* standard care): 0.1 (−7.0, 0.9) Effect estimate (95% CI), hydroxychloroquine *versus* standard care: −0.2 (−1.1, 0.6) | NR |
| Furtado *et al.* [44] | Hydroxychloroquine + AZ^a^ (n=214) | NR | Ventilation-free days: 0  (0, 14) | NR |
|  | Hydroxychloroquine^a^ (n=183) | NR | Ventilation-free days: 1  (0, 18) | NR |
|  | Between-group difference/comparison | NR | *P*=0.37 | NR |
| **Anti-inflammatory drugs** | | | | |
| Angus, DC *et al.* [53] | Hydrocortisone, fixed dose^b^ (n=137) | NR | NR | Respiratory support-free days, adjusted odds ratios: Mean (SD): 1.45 (0.34)  Median (95% CI): 1.42 (0.90, 2.24) |
|  | Hydrocortisone, shock-dependent dosing^b^ (n=141) | NR | NR | Respiratory support-free days, adjusted odds ratios: Mean (SD): 1.31 (0.30)  Median (95% CI): 1.28 (0.81, 2.00) |
|  | Standard care^b^ (n=101) | NR | NR | Respiratory support-free days, adjusted odds ratios: Mean: 1 (reference) Median: 1 (reference) |
|  | Between-group difference/comparison | NR | NR | Probability of superiority to no hydrocortisone, %: 94 (fixed dose), 85% (shock-dependent dosing) |
| Dequin, PF *et al.* [51] | Hydrocortisone (n=76) | NR | NR | Treatment failure on day 21^c^, %: 42.1 Endotracheal intubation (patients noninvasively ventilated at inclusion), %: 50  ECMO, %: 2.7 |
|  | Placebo (n=73) | NR | NR | Treatment failure on day 21^c^, %: 50.7 Endotracheal intubation (patients noninvasively ventilated at inclusion), %: 75 ECMO, %: 2.7 |
|  | Between-group difference/comparison | NR | NR | *P (treatment failure)*: 0.29 No comparison for other outcomes |
| **Kinase inhibitors** | | | | |
| Cao, Y *et al.* [54] | Ruxolitinib^a^ (n=20) | IMV: 0 | NR | NR |
|  | Placebo^a^ (n=21) | IMV: 5.0 (2.0, 8.0) | NR | NR |
|  | Between-group difference/comparison | NR | NR | NR |
| **Calcium release-activated calcium channel inhibitors** | | | | |
| Miller, J *et al.* [55] | Auxora^a^ (n=17) | NR | NR | Need for IMV or death within 30 days of randomisation: 17.6% |
|  | Standard care (n=9) | NR | NR | Need for IMV or death within 30 days of randomisation: 55.6% |
|  | Between-group difference/comparison | NR | NR | **HR: 0.23 (95% CI: 0.05, 0.96); *P*<0.05*** |
| **Anticoagulants** | | | | |
| Lemos, ACB *et al.* [56] | Therapeutic enoxaparin (n=10) | NR | NR | Median (IQR) ventilator-free days: 15 (6, 16) |
|  | LMW or unfractionated heparin (n=10) | NR | NR | Median (IQR) ventilator-free days: 0 (0, 11) |
|  | Between-group difference/comparison | NR | NR | **Median ventilator-free days: *P*=0.028* Ratio of successful liberation from mechanical ventilation: HR (95% CI): 4.0 (1.035, 15.053); *P*=0.031*** |
| **Repair therapies** | | | | |
| Cheng, L *et al.* [59] | rhG-CSF^a^ (n=100) | Oxygen: 10 (9, 12) | NR | NR |
|  | Standard care (n=100) | Oxygen: 10 (8, 13) | NR | NR |
|  | Between-group difference/comparison | Difference (95% CI): 0 (−1, 1) | NR | NR |

*: Denotes a statistically significant *P*-value or between-group comparison.

^a^: Treatment administered in addition to standard care, as defined by the investigators in each trial; ^b^: Participants could be randomly assigned to other interventions within other therapeutic domains; ^c^: Death or persistent dependence of mechanical ventilation or high-flow oxygen therapy.
AZ: azithromycin; CI: confidence interval; ECMO: extracorporeal membrane oxygenation; HR: hazard ratio; IFN: interferon; IMV: intensive/invasive mechanical ventilation; IQR: interquartile range: LMW: low molecular weight; NR: not reported; RBV: ribavirin; rhG-CSF: recombinant human granulocyte-colony stimulating factor; SD: standard deviation.

#### Supplementary Figure 1. PRISMA flow diagram of the SLR

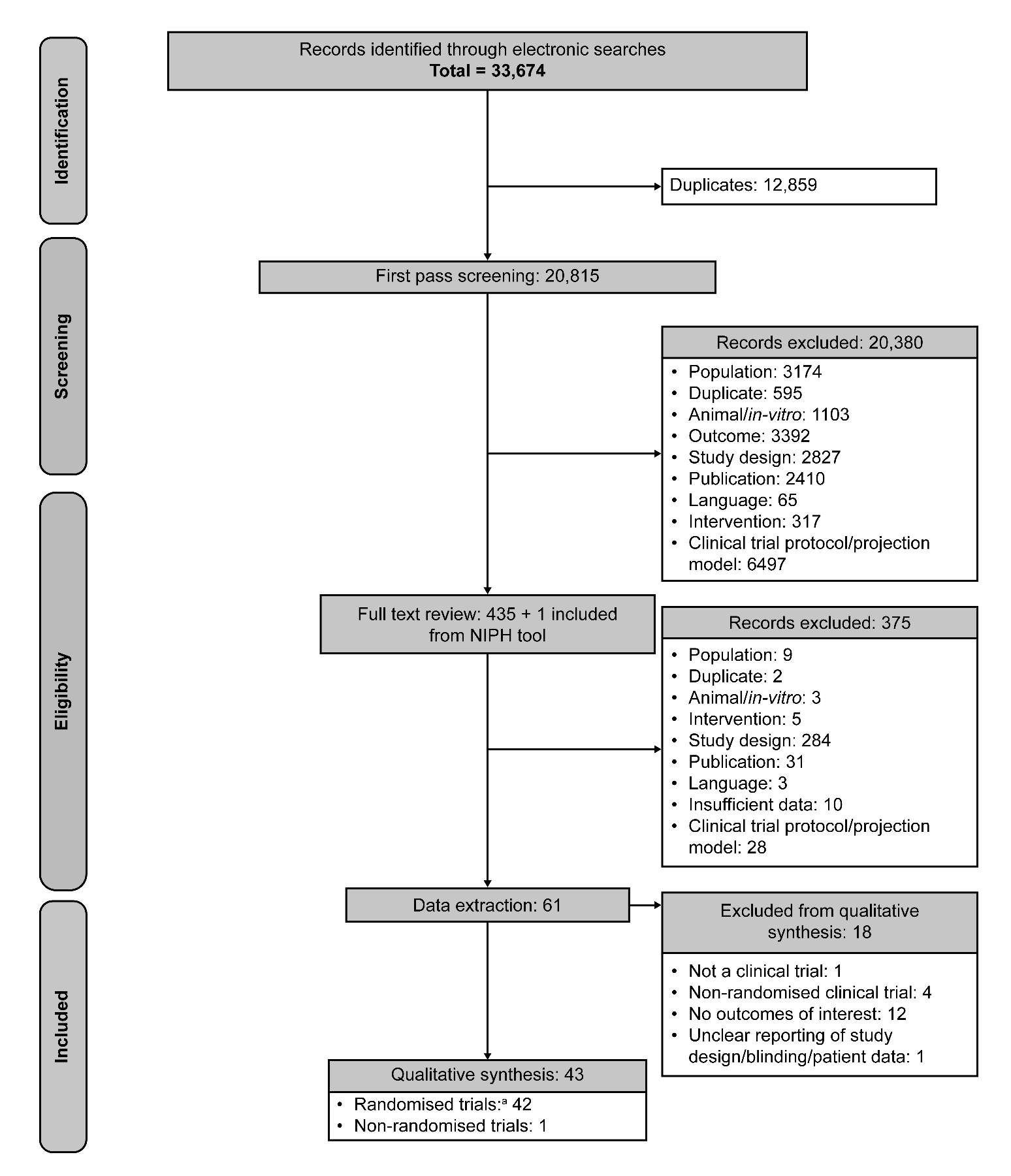

^a^: One study was a re-analysis of data from another included trial (Wang, Y *et al.*).
NIPH: Norwegian Institute of Public Health; PRISMA: Preferred Reporting Items for Systematic Reviews and Meta-Analyses; SLR: systematic literature review.

#### Supplementary Figure 2. Summary of studies included in qualitative synthesis in terms of study design (A), country (B), number of patients (C), hospitalisation status (D) and therapies assessed (E)

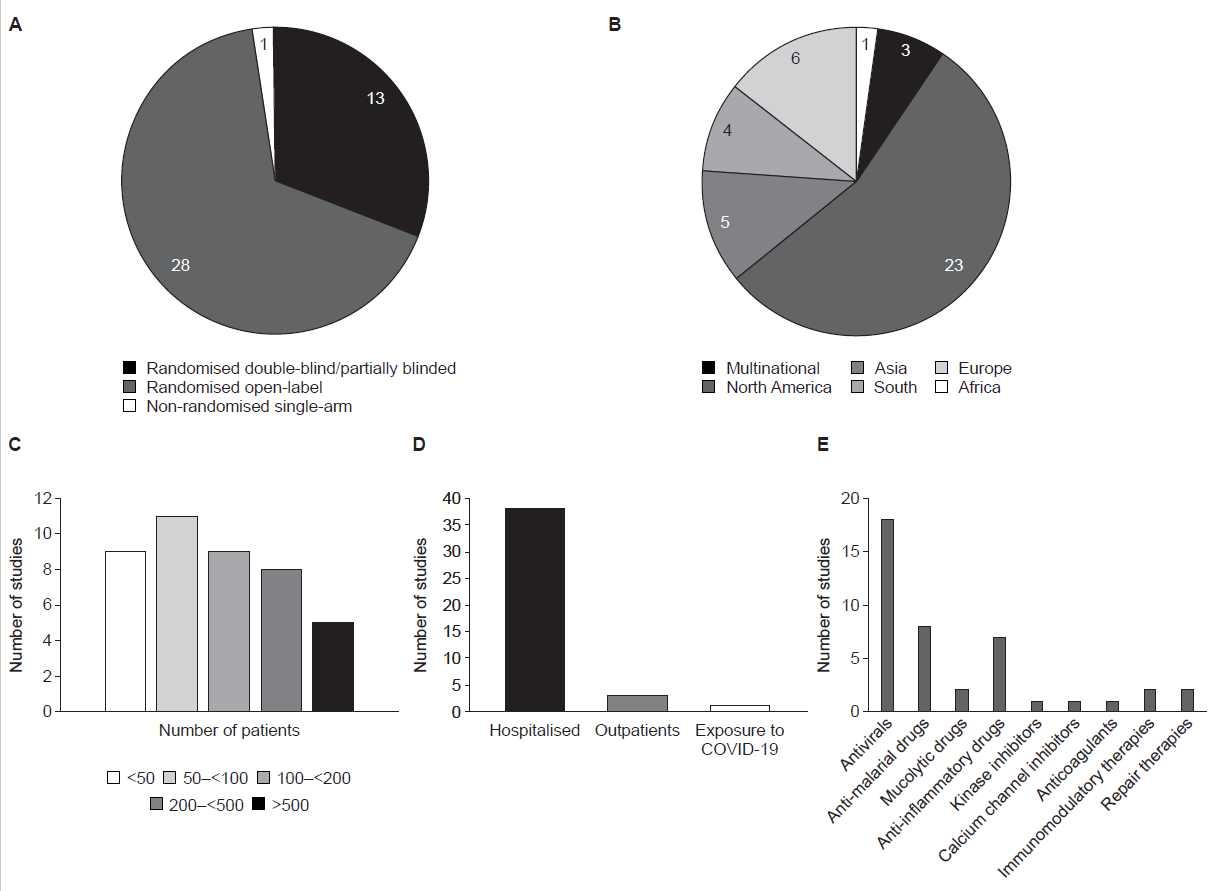
 Data presented do not include the article by Shih, WJ et al., as this was a re-analysis of data from another included trial (Wang, Y et al.).
COVID-19: Coronavirus disease 2019.

#### Supplementary Figure 3. Incidence of hospitalisation

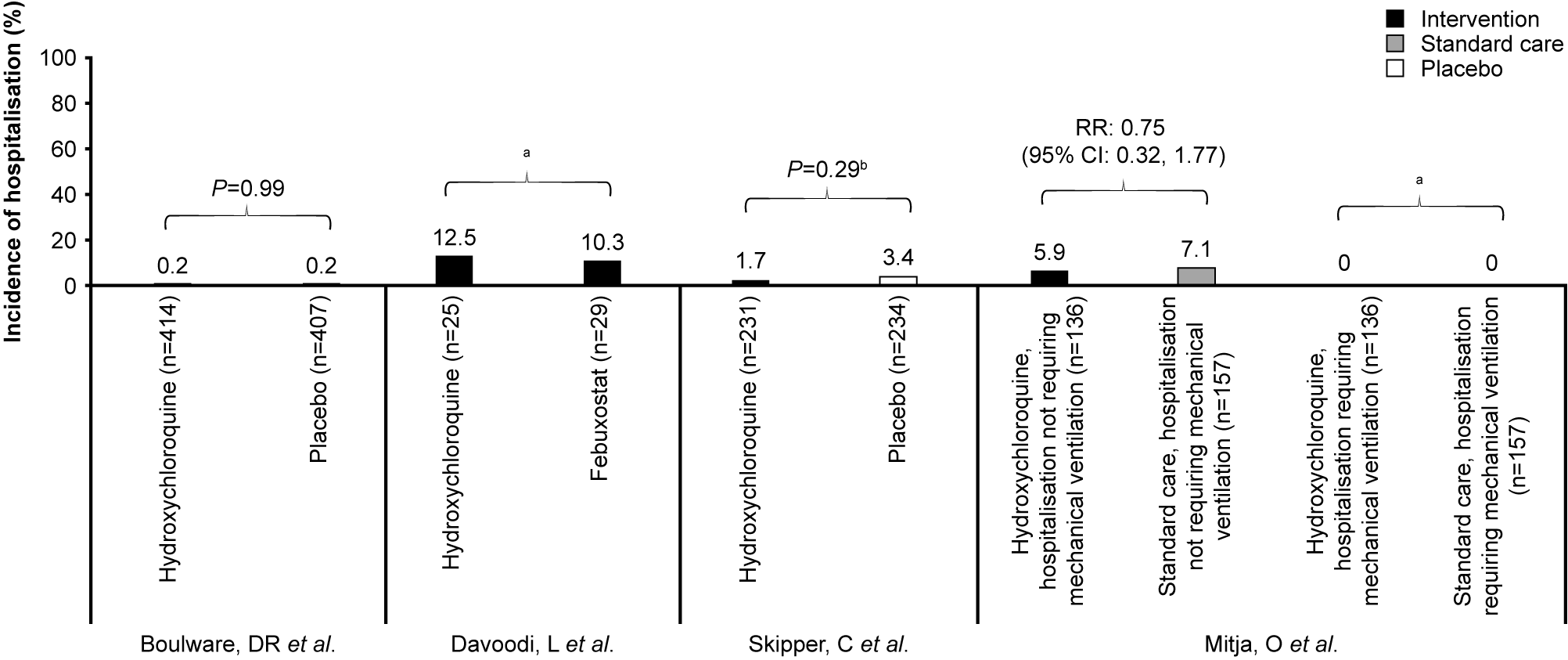

Data from trials reporting on non-hospitalised patients. *: Indicates a statistically significant *P*-value; ^a^: No between-group comparison; ^b^: *P*-value for the incidence of hospitalisation or death.
CI: confidence interval; RR: risk ratio.

#### Supplementary Figure 4. Quality assessment of trials included in qualitative synthesis

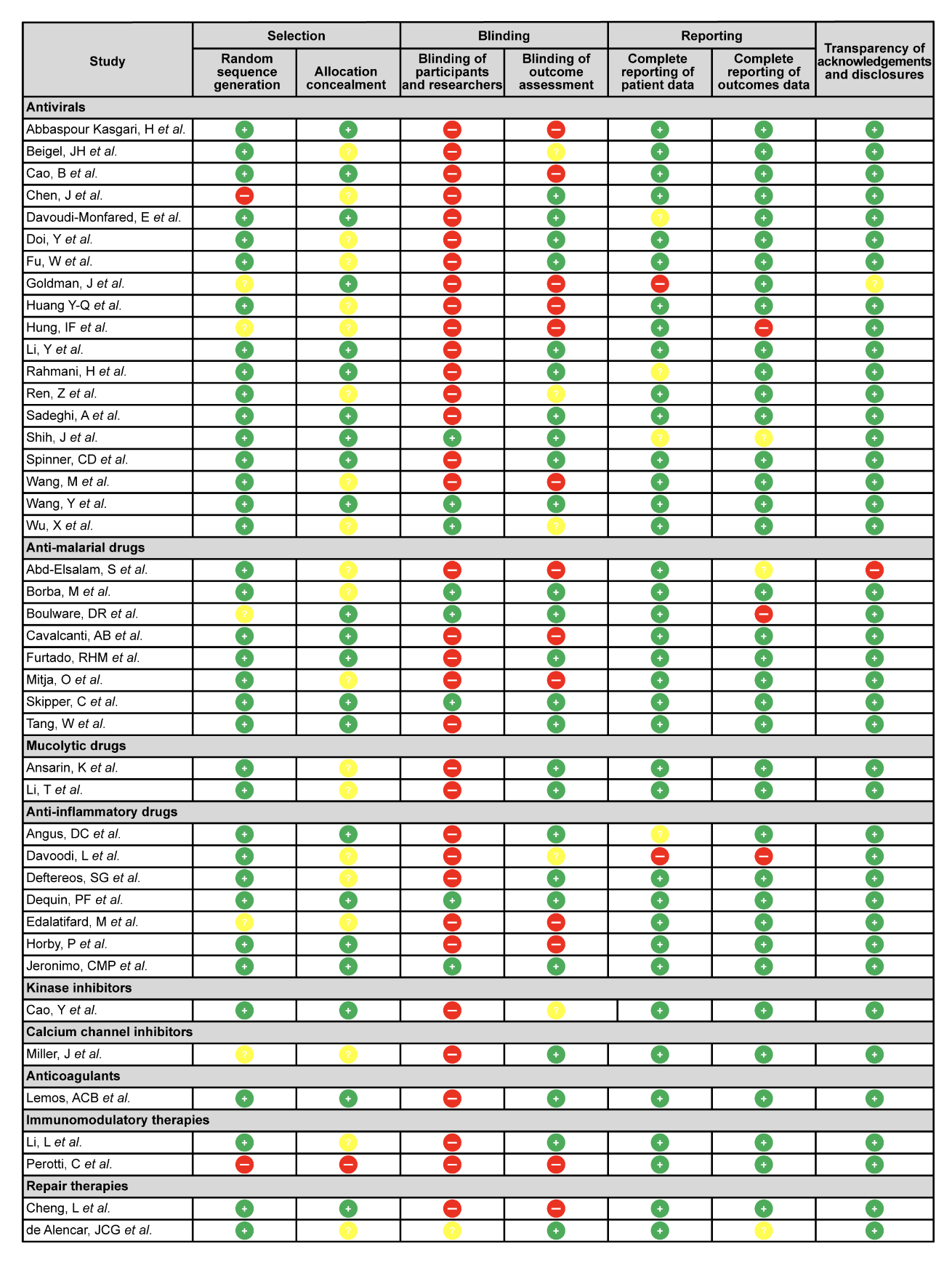

Red: high risk of bias; yellow: unclear risk of bias; green: low risk of bias.

Note: Shih, J *et al*. is a re-analysis of the study published by Wang, Y *et al*.
